# Fitness barriers to spread of colistin resistance overcome by first establishing niche in patients with enhanced colistin exposure

**DOI:** 10.1101/2021.06.11.21258758

**Authors:** Zena Lapp, Jennifer H Han, Divya Choudhary, Stuart Castaneda, Ali Pirani, Kevin Alby, Pam C Tolomeo, Ellie JC Goldstein, Ebbing Lautenbach, Evan S Snitkin

**Author notes:** **Corresponding author** Evan Snitkin, 1520D MSRB I, 1150 W. Medical Center Dr., Ann Arbor, MI, 48109-5680. **Author Contributions** E.S.S., J.H.H., E.L., and Z.L. conceptualized the study and acquired funding to support the project. K.A. performed MIC testing. Z.L, A.P., S.C., and D.C. performed data curation and formal analysis. Z.L. performed investigation and visualization. E.S.S. and Z.L. developed methodology. E.S.S. supervised the project. Z.L. and E.S.S. wrote the original draft. All authors reviewed and edited the manuscript. **Competing Interests Statement** J.H.H. is currently an employee of, and holds shares in, the GSK group of companies.

## Abstract

There is an urgent need to improve our understanding of how new antibiotic resistant organisms emerge and spread. A high-priority resistance threat is the ST258 lineage of carbapenem-resistant *Klebsiella pneumoniae*. Here, we studied resistance to the last-line drug colistin among ST258 by tracking its evolution across 21 U.S. hospitals over the course of a year. Phylogenetic analysis supported a significant fitness cost being associated with resistance, as resistance emergence was common but resistance variants were rarely transmitted. Furthermore, several resistance variants that were transmitted had acquired secondary variants that reverted the strain to susceptible. The exceptions to the general pattern of instability associated with resistance were two large clusters of resistant strains in one sublineage (clade IIB) present across Southern California hospitals. Quantification of transmission fitness in the healthcare environment indicated that, while resistant isolates from other clades were less fit than their susceptible counterparts, clade IIB resistant isolates were more fit, despite having similar resistance variants. Additional analyses supported the increased fitness of colistin-resistant clade IIB isolates being driven by a lineage-defining variant that increased clade IIB’s association with patient subpopulations with enhanced colistin exposure. These results show that a favorable genetic background and sustained selective pressure led to the emergence and spread of a colistin-resistant ST258 sublineage across a regional healthcare network. These findings highlight the utility of integrating pathogen genomic and corresponding clinical data from regional healthcare networks to detect emerging antibiotic resistance threats and understand the clinical practices and patient populations that drive their spread.

**Significance Statement:** Selective pressure in hospitals leads to frequent antibiotic resistance evolution. However, emergent resistance alleles are often not transmitted to other individuals because of fitness costs associated with resistance. Due to the difficulty of studying pathogen fitness in humans, our understanding of how resistant organisms circumvent these costs is limited. We integrate genomic and clinical data to understand the evolutionary trajectories leading to transmissible resistance for the last-line antibiotic colistin. While colistin resistance is generally associated with a fitness cost that hinders transmission, this cost was mitigated in a sublineage that had previously acquired mutations increasing its association with patient populations more likely to receive colistin, suggesting a key role for historical contingency in the emergence and spread of stable resistance.

## Introduction

Multidrug resistant organisms (MDROs) pose a significant threat to public health due to uncontrolled transmission and dwindling treatment options (1). Of greatest concern are epidemic MDRO lineages that have become adapted to healthcare settings and continually gain resistance due to selective antibiotic pressure (2, 3). As resistance evolution continues to outpace our ability to develop new therapies (3), there is a critical need to improve our understanding of the forces driving the emergence and spread of resistance so that we can maintain the efficacy of last-line antibiotics. While numerous studies have documented evolutionary trajectories leading to stable antibiotic resistance in laboratory settings (4–6), these findings may not translate to the real world due to a failure to capture relevant selective pressures associated with patient-to-patient transmission in healthcare settings (6). However, in part due to the difficulty of quantifying fitness effects outside of the lab, similar studies *in situ* are scarce.

When *de novo* evolution of antibiotic resistance occurs in an individual, there is often little to no subsequent transmission to others due to the fitness cost of maintaining resistance in the absence of antibiotic exposure (7, 8). However, in some cases this barrier is overcome, as evidenced by the proliferation of epidemic resistance clones (8, 9). This may happen when the resistance variant or gene has an inherently low fitness cost and is thus maintained in the population even in the absence of antibiotic exposure (10). Alternatively, the fitness cost of a given resistance element may depend on the genetic background in which it emerges, leading to dissemination of these elements only in certain lineages. Indeed, experimental evidence has shown that the genetic background of a strain interacts with resistance determinants to influence the fitness of the resistant strain (11, 12), highlighting the importance of historical evolutionary events in determining the potential ability of a resistant strain to spread. While the preferential spread of resistance elements in specific strains lends support to the importance of genetic background, in most instances it is unclear what the underlying epistatic interactions are that lead to these different fitness effects, particularly *in situ* (8). Understanding how the strain genetic background, in combination with selective pressures in healthcare settings, drives the emergence and spread of antibiotic resistance is critical for both early detection of high-risk clones and devising strategies to contain them.

Carbapenem-resistant *Klebsiella pneumoniae* (CRKP) is a global threat as it is resistant to nearly all available antibiotics and is associated with high mortality rates (1). One of the few remaining treatment options for CRKP is the antibiotic colistin (13). Concerningly, the viability of colistin as a treatment option moving forward is threatened by the frequent emergence of colistin resistance during treatment with this drug (14, 15). However, despite the accessibility of these resistance variants to CRKP, their spread to other patients is rare, with most cases even within a single healthcare setting stemming from parallel evolution (15). Thus, even though no fitness effects for colistin resistance have been detected *in vitro* (16–18), there appears to be a significant fitness cost *in situ*. Here, we document the emergence and spread of multiple colistin resistance variants in CRKP sequence type (ST) 258 across a regional healthcare network. Through integration of genomic and clinical data, we show how resistance spread was enhanced in genetic backgrounds that mitigated the fitness cost of resistance, thereby enabling regional dissemination of colistin-resistant lineages.

## Results

### Most resistant isolates contain variants in known resistance genes

Antimicrobial susceptibility testing revealed that 118/337 (35%) CRKP ST258 isolates were resistant to colistin, and that resistance was found consistently across long-term acute care hospitals (LTACHs) over the yearlong study (**Fig S1**). To identify likely resistance variants, we first searched all isolates for *mcr*-containing mobile genetic elements and variants in canonical chromosomal genes known to confer resistance (*pmrA, pmrB, phoP, phoQ, crrA, crrB*, and *mgrB*). We did not find *mcr* genes in any of the isolates, as expected since *mcr* genes have not been found in CRKP isolates from the U.S. during the timeframe of the study (July 2014 to August 2015) (19). However, the majority of colistin resistance we observed (103/118, 87%) could be explained by known and putative resistance variants in canonical resistance genes (**Fig S2**; **Table S1**; see methods for details on identification of resistance variants).

As not all resistance in our dataset could be explained by variants in known resistance genes, we next performed a genome-wide association study (GWAS)(20) on the isolates with unknown resistance determinants to identify putative non-canonical resistance-conferring variants. We identified two additional putative resistance genes in this way: *qseC* and phosphotransferase system sugar transporter subunit IIB (**Fig S2**; **Table S1**). These putative resistance variants account for 6/15 (40%) of the resistant isolates lacking variants in known resistance genes in our dataset. Notably, *qseC*, which explained resistance in 4 isolates, has been shown to confer colistin resistance in an experimental evolution study using clinical isolates (21). We were unable to identify putative colistin resistance-conferring variants for the other nine isolates. For all subsequent analyses, we defined resistance genes as the set of canonical and GWAS-identified resistance genes.

### Epistatic interactions appear to influence resistance in isolates with more than one variant in resistance genes

To further explore the relationship between genetic variation at putative resistance loci and colistin resistance we next examined the number of variants in resistance genes among resistant and susceptible isolates. Supporting a causal role for genetic variation in putative resistance loci, we observed that while only 9/154 (5.8%) isolates without a variant in a putative resistance locus were phenotypically resistant, 93/122 (76.2%) isolates with a single variant in a putative resistance locus were colistin resistant. However, when expanding to look at isolates with two or more variants in putative resistance loci, the association with resistance inverted, with 45/61 (73.8%) isolates being phenotypically susceptible and 16/61 (26.2%) resistant (**Fig 1A**). This unexpected relationship between accumulation of multiple resistance variants and susceptibility was also observed at the level of colistin MIC, with susceptible isolates with two or more variants in resistance genes having lower MICs than those with one or fewer (**Fig 1B**; median MIC of 0.5 vs. 1, Wilcox p = 0.0002).

**Figure 1:**
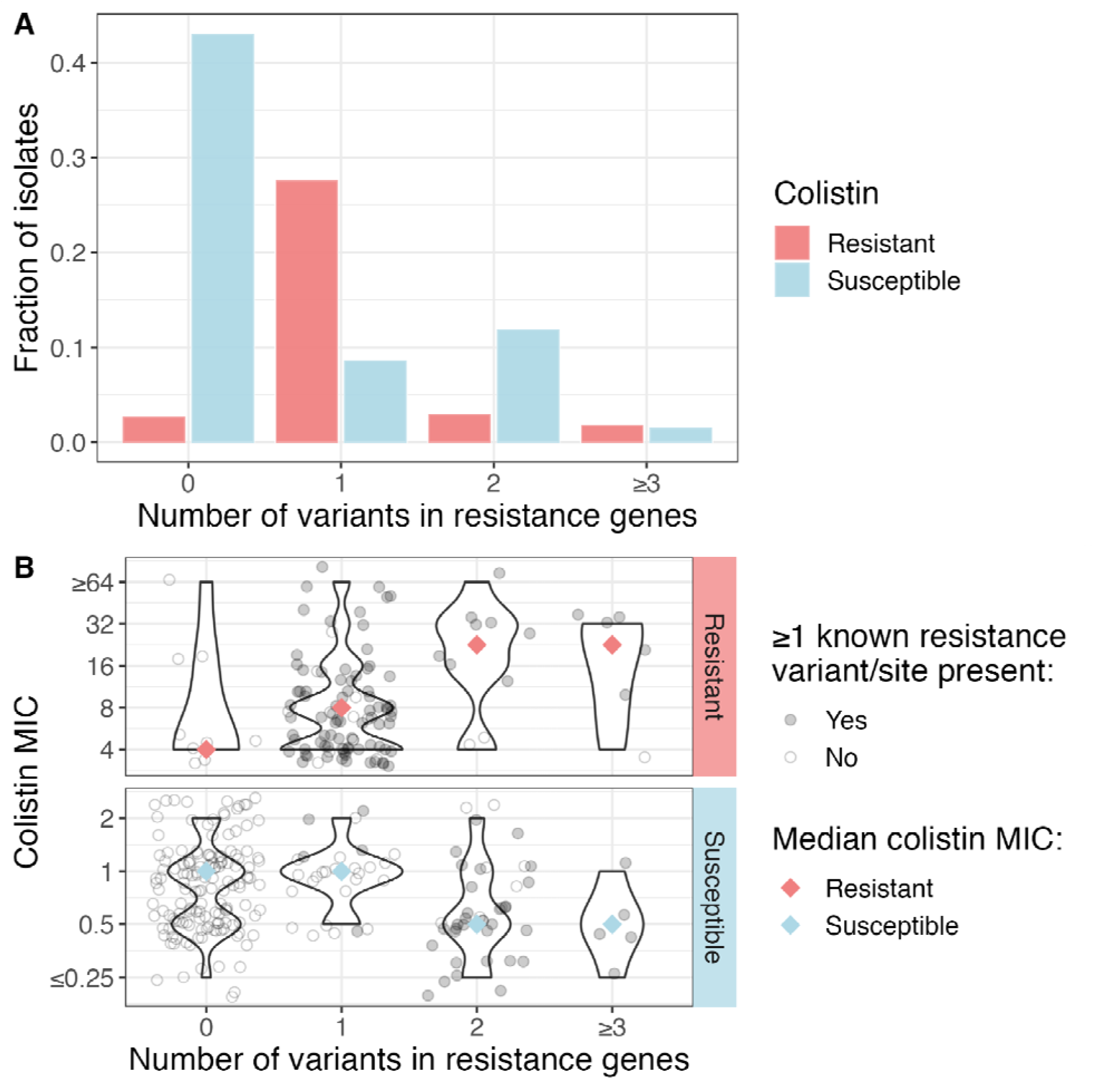
Epistatic interactions appear to influence the extent of colistin resistance in clinical CRKP isolates. (A) Most susceptible isolates have no variants in known resistance genes, while most resistant isolates have at least one variant in a known resistance gene. Many susceptible isolates also have two variants in known resistance genes, suggesting potential reversion events. (B) Colistin MIC split by total number of variants in resistance genes. Shapes indicate whether any of the variants are putative resistance variants (see methods for definition of resistance variants). On average, resistant isolates with two or more variants in resistance genes have a higher MIC than resistance isolates with one or fewer variants in resistance genes, and susceptible isolates with two or more variants in resistance genes have a lower MIC than susceptible isolates with one or fewer variants in resistance genes. Significance defined as Wilcox p < 0.05. MIC=minimum inhibitory concentration.

We hypothesized that the enrichment for susceptibility among isolates with multiple variants in resistance loci could be a consequence of the fitness cost of resistance leading to selection for secondary mutations that abrogated resistance, and thereby mitigated the fitness cost. Supporting this hypothesis, we found that in 36/45 (80%) cases where susceptible isolates harbored multiple variants in resistance genes, at least one of the variants or variant sites had been previously shown to confer resistance *in vitro* (**Table S1**), suggesting that the secondary variants present in resistance genes may be suppressor mutations that revert historically resistant strains back to susceptible. This hypothesis was further corroborated by visualizing these variants on the phylogeny (**Fig 2**). For instance, we observed several instances where putative suppressor mutations have arisen in a lineage containing a resistance-conferring *phoQ* mutation. These and other putative suppressor mutations usually occurred within the same gene or in a downstream gene of the molecular pathway to resistance (**Fig S3**), and were sometimes followed by re-acquisition of resistance through a different mechanism (**Fig 2**; **Fig S4**). Taken together, these findings suggest that, while variants in resistance genes are often associated with an increase in MIC (**Fig 1B**), there also exist epistatic interactions among variants in these genes that may lead to colistin susceptibility, even in strains containing known resistance-conferring mutations (22, 23).

**Figure 2:**
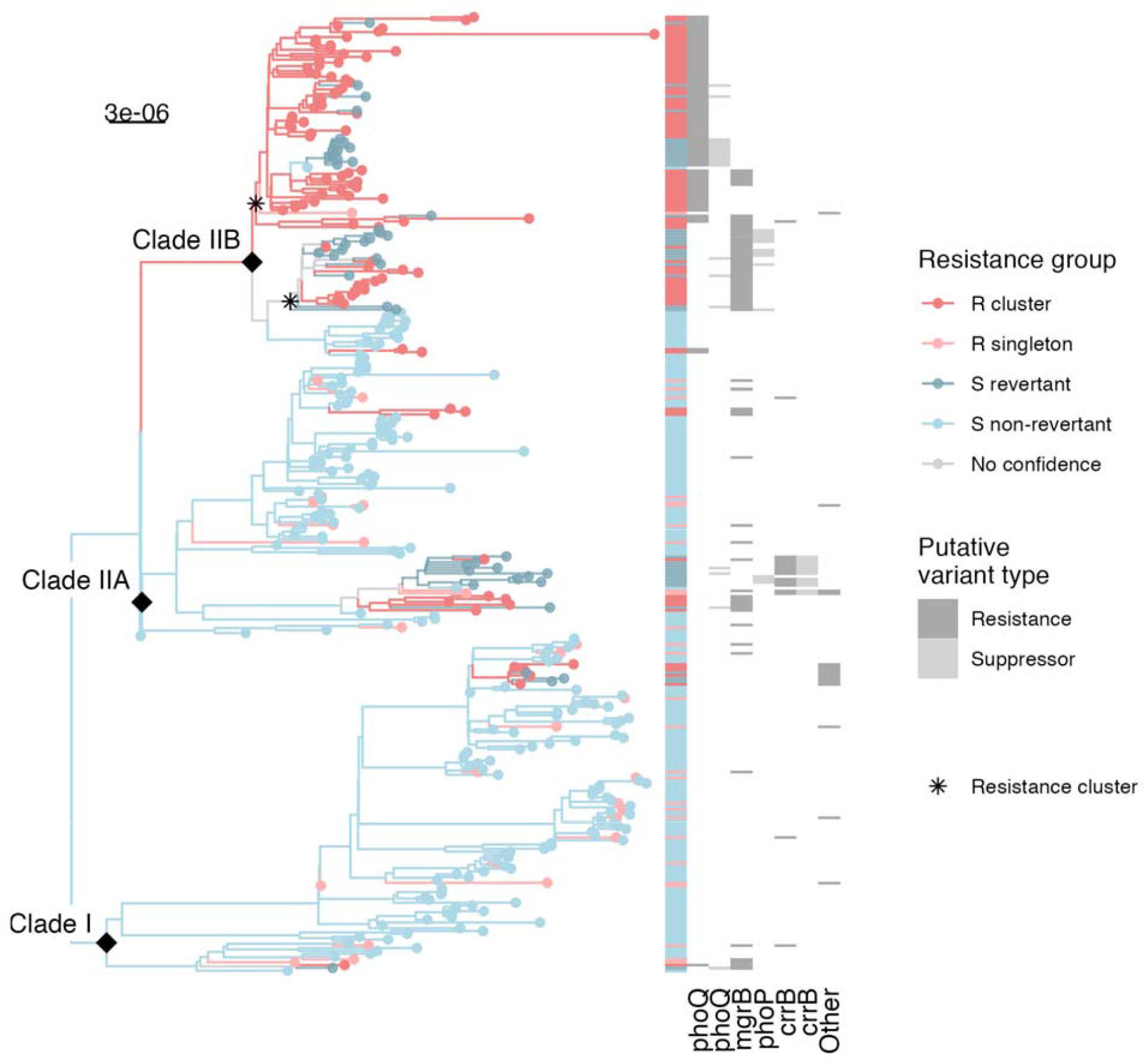
Distinct resistance evolution patterns exist in different CRKP ST258 sublineages. Clade IIB exhibits extensive dissemination of resistance variants, as well as several putative reversion events, while the other clades contain sporadic and less transmitted resistance variants. Each grey column of the heatmap is a gene, where all variants of a given type (resistance in dark grey, suppressor in light grey), are grouped for clarity. See Fig S5 for more details on individual variants. The asterisks indicate the 2 clade IIB resistance clusters mentioned in the text. Scale bar is in substitutions per site per year. R=resistant; S=susceptible.

### Colistin resistance exhibits patterns of *de novo* evolution, onward dissemination, and reversion to susceptibility

To better understand the origin and fate of colistin resistance variants, we further investigated the phylogenetic relationship among colistin-resistant and susceptible isolates (**Fig 2**; **Fig S5**). Based on the observed resistance distribution on the phylogeny we classified each isolate into one of four groups: resistant cluster (sub-clade harboring multiple resistant isolates), resistant singleton (single resistant isolate with susceptible phylogenetic neighbors), susceptible revertant (susceptible but contains a putative resistance-conferring variant), and susceptible non-revertant (other susceptible isolates). Grouping resistance in this way revealed that, while 70% (28/40) of resistance emergence events did not spread between patients (i.e. were resistant singletons), the 12 that did spread accounted for 76% (90/118) of resistance across study facilities. Visualization of these resistance clusters on the phylogeny revealed a striking dichotomy between a clade II sublineage (referred to as clade IIB) and the other ST258 sublineages (clades I and IIA). Clade IIB contained two large clusters of colistin-resistant isolates, while the other two sublineages mainly exhibited sporadic parallel evolution of resistance. In total, these two clonal expansions in clade IIB, which occurred across 11/12 LTACHs in Southern California (10 in the Los Angeles area and 1 in San Diego; **Fig S6**), accounted for over half (69/118, 58%) of resistance in the isolate collection. Each cluster harbored a distinct resistance mutation, with one being associated with a nonsense mutation in *mgrB* (Gln30*) and the other a missense mutation in *phoQ* (Thr244Asn), respectively. Notably, these same types of variants occurred in the sporadic colistin-resistant isolates (**Fig 2**; **Fig S4**), suggesting that these specific resistance variants were likely not inherently more fit. Within these clonal expansions, many of the isolates appeared to regain susceptibility to colistin via the accumulation of additional variants in resistance genes as described above, suggesting that despite their proliferation, there is still a fitness cost associated with resistance.

### Resistant strains in clade IIB are more fit than their susceptible non-revertant counterparts

Next, we were interested in understanding whether the widespread dissemination of colistin resistance variants in clade IIB could be explained by an increased ability to spread relative to resistant strains in the other two clades. In particular, we hypothesized that the success of resistance variants in clade IIB could be a consequence of a decreased fitness cost associated with resistance, as compared to the cost of resistance in other clades. While previous studies have not found a fitness cost associated with colistin resistance in *K. pneumoniae* (16–18), this is inconsistent with observed lack of onward transmission of most resistant mutants (15). We hypothesized that this was because the measurement of fitness in laboratory conditions fails to capture the range of stresses encountered during colonization and transmission among patients in healthcare settings. To overcome this issue and estimate *in situ* transmission fitness of resistant and susceptible isolates from different lineages in the context of the healthcare environment, we used time-scaled haplotypic density (THD), an analytic approach that uses the genetic relatedness between isolates to quantify the extent of relative spread of each strain (24). A higher THD value is consistent with higher transmission fitness (i.e. ability to spread).

In clades I and IIA we observed evidence of a significant transmission fitness cost for resistance variants, as resistance clusters and susceptible revertants were less able to spread than susceptible non-revertants from those clades (**Fig 3**; R cluster: Wilcox p = 0.04; S revertant: Wilcox p = 0.02).. In contrast to clades I and IIA, resistance variants in clade IIB were associated with a significant transmission fitness benefit, with resistant clusters and susceptible revertants from clade IIB being more able to spread than susceptible non-revertants from that clade (R cluster: Wilcox p = 0.03; S revertant: Wilcox p = 0.009). Taken together, these findings suggest that resistance variants in isolates from clade IIB conferred a transmission fitness advantage, which may account for the independent emergence and spread of two different resistance alleles in this clade. In contrast, the accumulation of variants in resistance genes in the other clades appears to be associated with a transmission fitness cost, which is consistent with their limited clonal spread.

**Figure 3:**
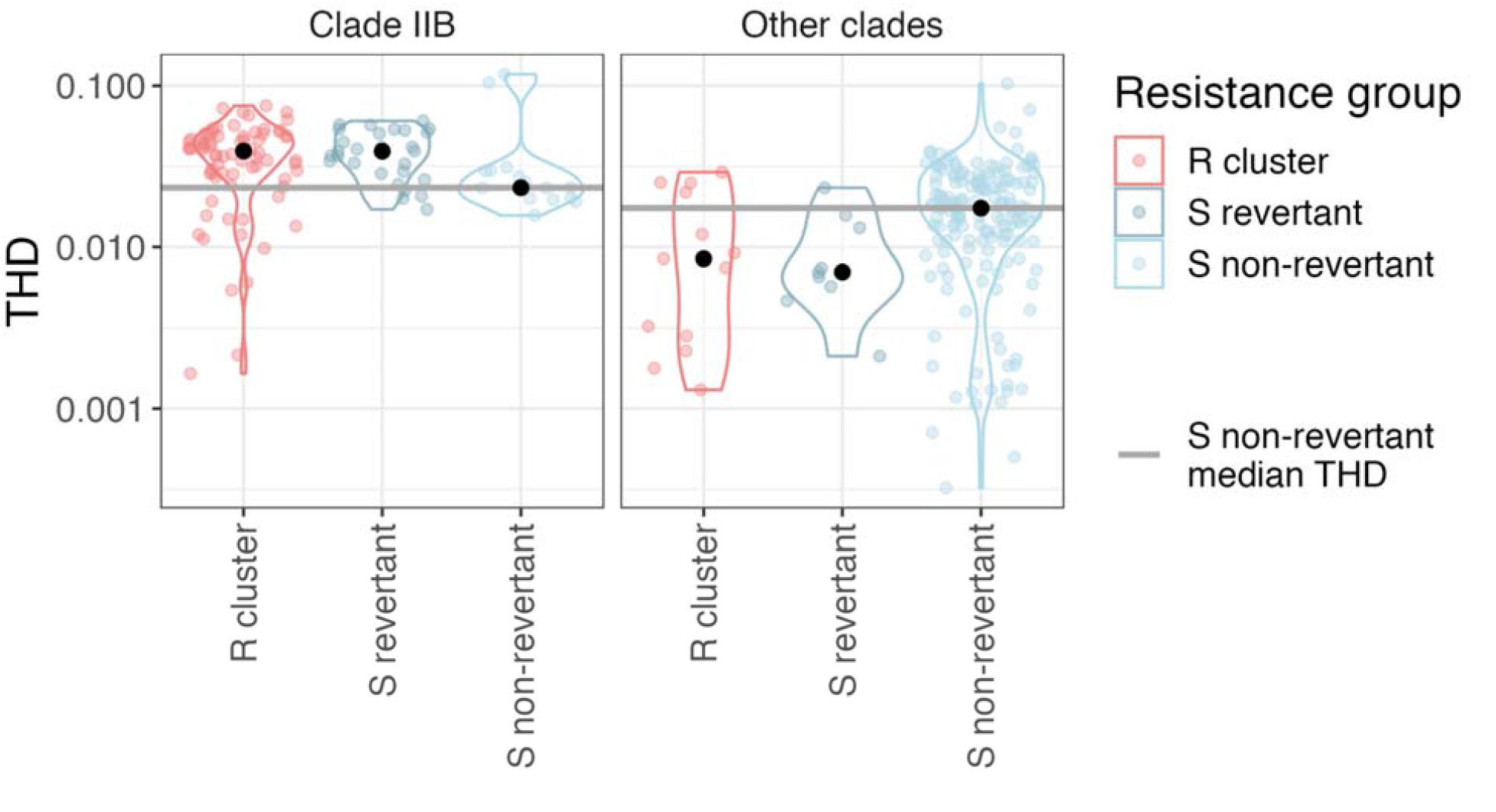
Colistin resistance does not impart a fitness cost in clade IIB strains. In clade IIB, resistant isolates are more fit than corresponding susceptible non-revertants. On the other hand, resistant isolates in other clades are less fit than susceptible non-revertants in those clades. Significance defined as Wilcox p < 0.05. R=resistant; S=susceptible; THD=time-scaled haplotypic density.

### Clade IIB isolates have more colistin exposure compared to isolates from other clades

Lastly, we investigated whether colistin use in study facilities influenced the emergence and spread of stable resistance in clade IIB. Supporting the role of colistin use in the spread of colistin resistance, we found a positive association between having a clade IIB isolate and both prior exposure to colistin (15/120, 12.5% in clade IIB vs. 8/217, 3.7% in other clades; Fisher’s exact p = 0.003), as well as treatment with colistin (41/120, 34.2% in clade IIB vs. 39/217, 18.0% in other clades; Fisher’s exact p = 0.001), regardless of colistin mutation status. It is notable that most of the colistin exposure in patients harboring clade IIB isolates occurred after patients acquired their isolates, which suggests that the association isn’t simply a reflection of antibiotic exposure increasing the risk for acquiring isolates resistant to the cognate antibiotic. Rather, we hypothesized that this association between colistin use and clade IIB is mediated by our previously reported observation that clade IIB is enriched in respiratory isolates (25) and the frequent use of colistin to treat respiratory tract infections during the study period (**Fig 4A**). This hypothesis is supported by a slight enrichment in colistin treatment for clade IIB respiratory isolates compared to non-respiratory isolates (31/83, 37.3% for respiratory vs. 10/37, 27.0% for non-respiratory; Fisher’s exact p = 0.3), and a significant enrichment across all clades (53/183, 29% for respiratory vs. 27/154, 17.5% for non-respiratory; Fisher’s exact p = 0.01). Furthermore, in addition to potentially influencing the spread of colistin resistance, the extensive use of colistin to treat infections in a setting of clonally disseminating colistin resistance led to 36/118 (30.5%) patients with colistin-resistant isolates being treated with this antibiotic, even though the treatment was likely ineffective (**Fig 4B**).

**Figure 4:**
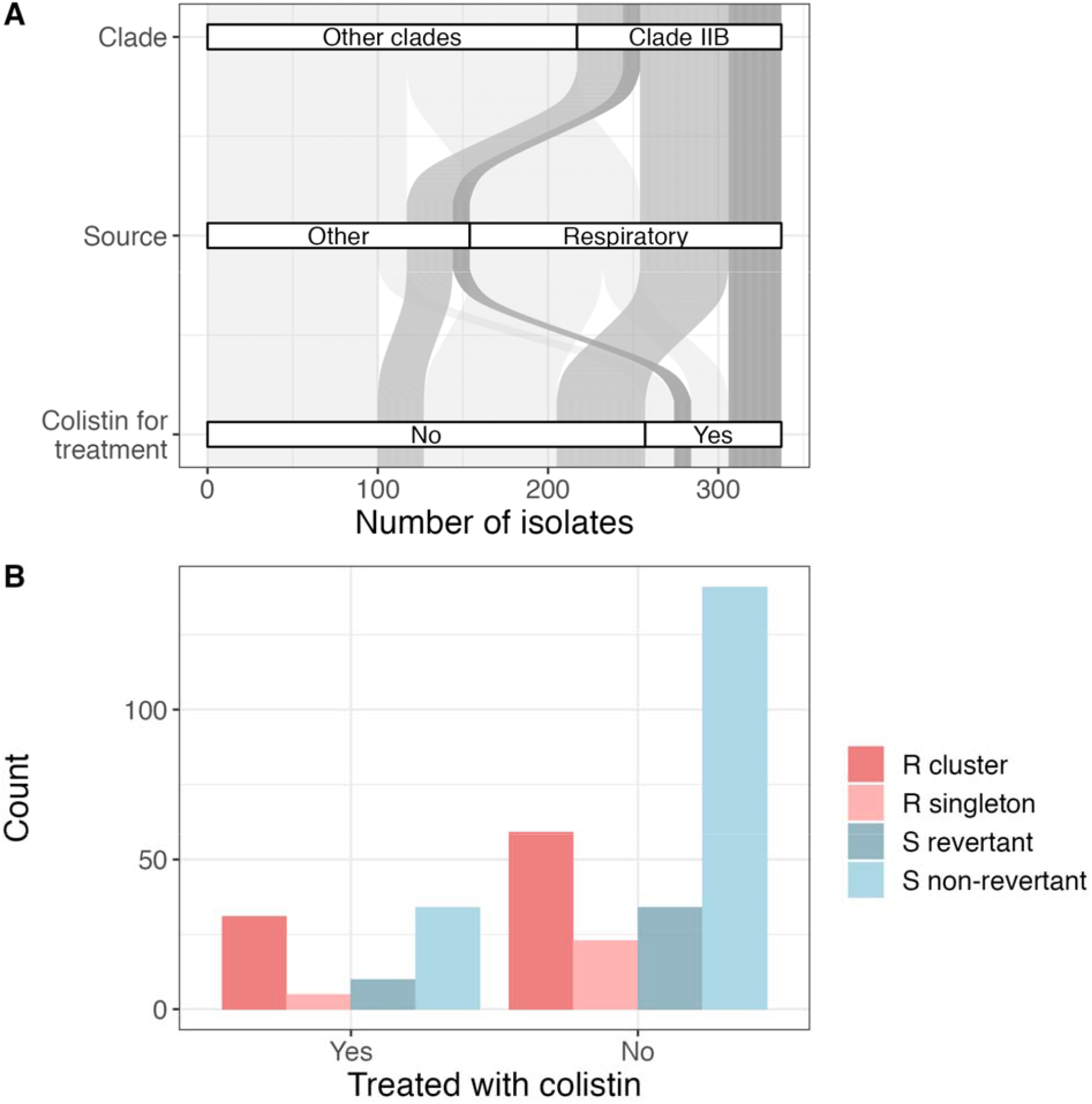
Patients with colistin-resistant isolates were treated with colistin. (A) The majority of clade IIB isolates are respiratory isolates, and many of the patients with those strains were treated with colistin (empiric and/or definitive). Differences in transparency are aesthetic to draw attention to clade IIB isolates treated with colistin. (B) Many colistin-resistant isolates were treated with colistin.

In putting our past and present observations together, we noted that a defining feature of clade IIB was disruption of the putative O-antigen glycosyltransferase *kfoC* (25). Therefore, we hypothesize that disruption of *kfoC* increased the affinity of clade IIB for the respiratory tract, which in turn resulted in the clade IIB lineage having increased rates of colistin exposure, and thereby decreased the transmission fitness cost of colistin resistance, allowing colistin-resistant clade IIB strains to spread across a regional healthcare network (**Fig 5**).

**Figure 5:**
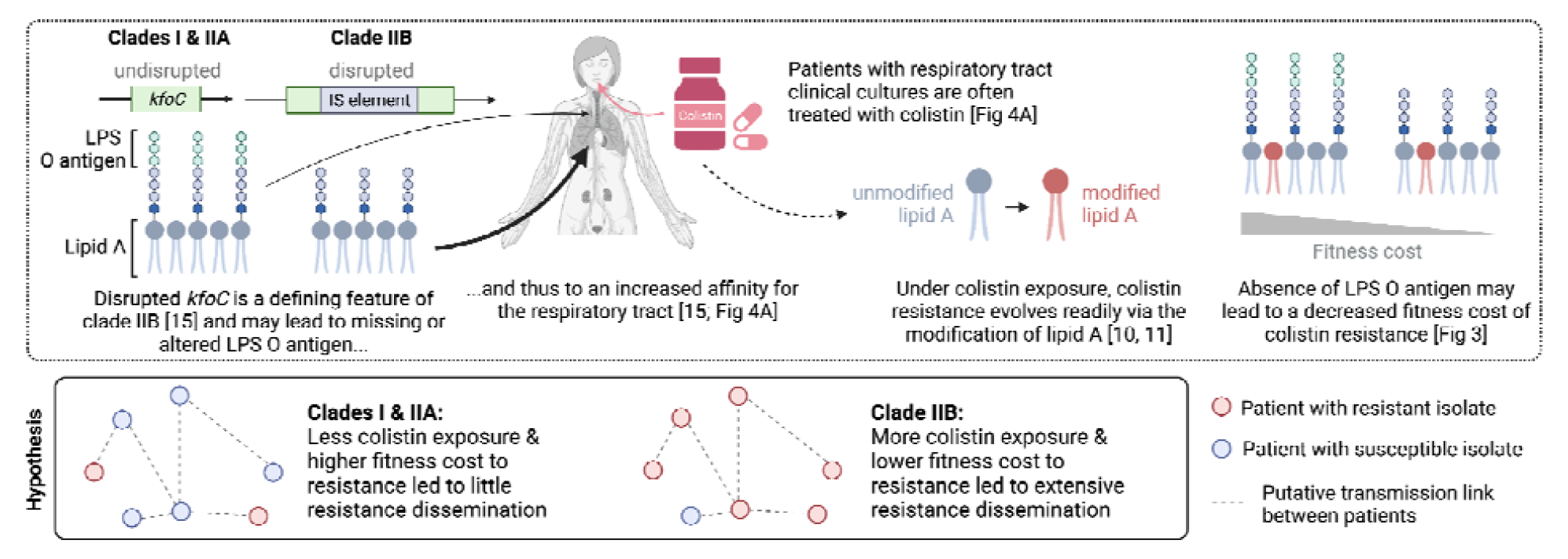
Disruption of the putative O-antigen glycosyltransferase *kfoC* may lead to decreased transmission fitness cost of colistin resistance. Created with BioRender.com

## Discussion

Improving our understanding of how antibiotic resistance evolves and disseminates in the healthcare setting is critical for optimizing surveillance, antibiotic stewardship, and infection prevention strategies. Here, we used genomic data from a comprehensive regional sample of clinical CRKP ST258 isolates to track the origin and fate of colistin resistance variants, and then overlaid clinical data to elucidate the drivers of resistance spread. Our analysis identified a circulating sublineage whose genetic background appears to decrease the fitness cost of resistance *in situ* by increasing association with patient populations with increased colistin exposure, thus allowing colistin-resistant strains to spread among regional healthcare facilities. More broadly, these findings highlight how regional genomic surveillance can be applied to hone in on emerging resistance threats with epidemic potential, as well as uncover patient populations and antibiotic usage patterns accelerating their spread.

Examination of colistin resistance variants within the comprehensive and longitudinal context of circulating CRKP strains allowed for inferences into their functional impact and clinical significance. We discovered that resistant isolates with multiple variants in resistance genes have increased resistance, but susceptible isolates with multiple variants in resistance genes have decreased resistance. Additionally, we identified several instances where a known resistance variant was present in susceptible isolates, likely due to a suppressor variant in either the same resistance gene or a downstream resistance gene (22, 23). This finding has two important implications. First, known resistance variants occurring in both resistant and susceptible isolates complicates the computational identification of novel resistance determinants using GWAS. Possible solutions include increasing sample size or removing isolates with known resistance variants. Second, the presence of a colistin resistance variant cannot necessarily be equated with *in vitro* phenotypic resistance. Clinically, this suggests that testing for resistance variants may not be a substitute for testing for phenotypic resistance, as colistin treatment may still be effective for some strains harboring resistance variants. In addition to being phenotypically susceptible, revertants that harbor resistance variants may in general have more difficulty becoming resistant once more, which may lend credence to treating these patients with colistin (23).

Of particular genomic and clinical interest are two independent instances of emergence and spread of colistin resistance variants in clade IIB, in stark contrast to the sporadic emergence of resistance variants in the other clades. Notably, the types of resistance variants present in clade IIB also occurred in other clades, implying that the nature of the variant itself is not what allowed the clade IIB variants to spread. Instead, our analysis of transmission fitness using THD suggests that these colistin resistance variants were able to spread in this clade because the genetic background of the clade reduced the fitness cost of the variants. We previously showed that a defining feature of clade IIB is that the majority of isolates contain a disruption in the lipopolysaccharide (LPS) O locus gene *kfoC* (25), a putative glycosyltransferase. Given the key role that LPS plays in both colistin-mediated killing and colistin resistance (14), it is possible that inactivation of *kfoC* facilitates the spread of colistin resistance by altering the outer membrane in a way that reduces the fitness cost of resistance-conferring LPS modifications. Furthermore, our data supports that *kfoC* disruption may indirectly mitigate the fitness cost of colistin resistance by increasing the association between clade IIB and patient populations in which colistin use is more frequent. In particular, clade IIB isolates are significantly enriched in both association with the respiratory tract and colistin exposure. Thus, increased colistin use in clade IIB may have both set the stage for the emergence of multiple instances of stable resistance in this clade and provided a continued selective pressure to maintain resistance. These findings highlight the importance of continued real-time monitoring of resistance to improve antibiotic stewardship and decrease the likely ineffective use of colistin to treat colistin-resistant CRKP strains.

One strength of our study is that we have comprehensive sampling of clinical CRKP isolates from LTACHs over the course of a year. This not only allowed us to capture putative transmission events of colistin resistance, but also allowed us to estimate the transmission fitness of different ST258 sublineages *in situ*. Additionally, whole-genome sequencing of study isolates permitted us to interrogate not only variants in known resistance genes, but also investigate putative resistance-conferring variants in other genes using GWAS. Furthermore, our access to clinical information about colistin use allowed us to identify a relationship between this clinical variable and different ST258 sublineages, providing us with insight into the selective pressures of colistin resistance evolution and spread in patients.

Our study also has several limitations. First, we do not have rectal surveillance cultures, which limits our ability to fully capture the population of colistin-resistant and susceptible CRKP in the LTACHs studied, and thus to fully define spread. While this could bias our clinical collection if certain strains are more likely to present clinically and be cultured at extraintestinal sites (e.g. clade IIB vs. other clades), the strains we have included are the most clinically relevant since they were identified in clinically derived samples. Also, we cannot be certain that our classification of isolates into different categories of colistin susceptibility and resistance is entirely accurate. For instance, putative *de novo* evolution of resistance in resistant singletons may not have occurred in the patient we sampled. However, our comprehensive sampling of clinical isolates from the facilities in the study provides some confidence that resistance in resistant singleton isolates did in fact evolve in that patient. Another limitation of our study is that we do not have information about colistin exposure prior to the patient entering the healthcare facility, and we do not have information about the dose or duration of colistin use. Even with this limitation, we were able to identify significant associations between colistin use and different ST258 sublineages.

In conclusion, we identified an emerging ST258 sublineage that is more fit and more amenable to maintaining colistin resistance than other ST258 strains *in situ*, likely due to an altered genetic background. This is of particular concern due to the already limited treatment options for CRKP, and therefore merits further surveillance to determine the extent of spread. Furthermore, our findings highlight how the inclusion of relevant clinical data amplifies the value of regional genomic surveillance by enabling identification of novel resistance and virulence variants of interest, as well as revealing the clinical practices and patient populations driving their spread.

## Materials and Methods

### Study isolates and metadata

We used whole-genome sequences of clinical CRKP isolates from a prospective longitudinal study in 21 U.S. LTACHs over the course of a year (BioProject accession no. <u>PRJNA415194</u>) (26). Isolates and metadata were collected, and isolates were sequenced, as described in Han *et al*. (26). Prior patient use of colistin in the facility from which the isolate was taken (up to 30 days before the isolate collection date), as well as colistin use for empiric and definitive treatment, was extracted from the electronic health record. We only used information on colistin, not polymyxin B, for all analyses.

### Antibiotic susceptibility testing

Custom Sensititre broth microdilution panels (ThermoFisher Scientific, Waltham, MA) were used to determine colistin MICs. We used Clinical & Laboratory Standards Institute (CLSI) guidelines for interpretations with an MIC cutoff of ≥ 4 characterized as resistant (27). CLSI guidelines classify isolates with an MIC ≤2 as being of intermediate resistance and therefore are technically considered non-resistant using this interpretation; however, the European Committee on Antimicrobial Susceptibility Testing defines isolates with an MIC ≤2 as susceptible (28). For clarity, we chose to use the term susceptible for isolates with an MIC ≤2.

### Isolate selection

Multi-locus sequence types were called using ARIBA (29). Over 90% of CRKP isolates collected over the course of the study belonged to ST258; therefore, we focus all of our analyses on this sequence type. We ordered the isolates by resistance status followed by collection date. For all analyses except GWAS, only the first patient ST258 isolate from this ordered list was used, thus prioritizing resistant isolates over susceptible isolates. Sample sizes were too small to glean insights into within-host colistin resistance evolution.

### Single nucleotide variant calling, indel calling, and phylogenetic tree reconstruction

Variant calling was performed with a customized variant calling pipeline (https://github.com/Snitkin-Lab-Umich/variant_calling_pipeline/) as follows. The quality of sequencing reads was assessed with FastQC v0.11.9 (30), and Trimmomatic v0.39 (31) was used for trimming adapter sequences and low-quality bases. Single nucleotide variants (SNVs) were identified by (i) mapping filtered reads to the ST258 KPNIH1 reference genome (BioProject accession no. PRJNA73191) using the Burrows-Wheeler short-read aligner (bwa v0.7.17) (32), (ii) discarding polymerase chain reaction duplicates with Picard v2.24.1 (33), and calling variants with SAMtools and bcftools v1.9 (34). Variants were filtered from raw results using VariantFiltration from GATK v4.1.9.0 (35) (QUAL >100; MQ >50; ≥ 10 reads supporting variant; and FQ < 0.025). Indels were called using the GATK HaplotypeCaller (36) with the following filters: root mean square quality (MQ) > 50.0, GATK QualbyDepth (QD) > 2.0, read depth (DP) > 9.0, and allele frequency (AF) > 0.9. In addition, a custom Python script was used to filter out (mask) variants in the whole-genome alignment that were: (i) SNVs <5 base pairs (bp) in proximity to indels, (ii) in a recombinant region identified by Gubbins v2.3.4 (37), in a phage region identified by the Phaster web tool (38) or (iii) they resided in tandem repeats of length greater than 20bp as determined using the exact-tandem program in MUMmer v3.23 (39). This whole-genome masked variant alignment was used to reconstruct a maximum likelihood phylogeny with IQ-TREE v1.6.12 (40) using the general time reversible model GTR+G and ultrafast bootstrap with 1000 replicates (-bb 1000) (41).

### Insertion calling

Large insertions relative to the reference genome were called using panISa v0.1.4 (42).

### Variant preprocessing

We preprocessed variants to include multiallelic sites and used the major allele method for variant binarization, as described in Saund *et al*. (43). We used SnpEff to predict the functional impact of single nucleotide variants and indels (high, moderate, low, modifier) (44). Additionally, we considered all insertions in or upstream of genes as high impact, and those downstream of genes as moderate impact. Only high and moderate impact variants were included in downstream analyses.

### Identification of putative resistance variants

#### Known resistance genes

We consider the following genes known (canonical) resistance genes: *mgrB, phoP, phoQ, crrA, crrB, pmrA*, and *pmrB* (45). We group variants in known resistance genes into the following categories, in order of decreasing confidence:

1. <u>Known:</u> experimentally confirmed resistance variants, including loss-of-function variants in *mgrB* (45–48).
2. <u>Known site:</u> the variant occurs at a nucleotide site where there is an experimentally confirmed resistance variant, but that specific amino acid change has not been experimentally confirmed (22, 49–51).
3. <u>Putative:</u> nonsynonymous or disruptive variants in known resistance genes where >60% of the variants are present in resistant isolates.

#### Genome-wide association study

We performed a burden test using treeWAS v1.0 (20), a convergence-based GWAS method, as our sample size is relatively small and resistance is a very convergent phenotype. A burden test increases power to detect resistance genes, and convergence-based methods control for the structure of the phylogeny more than mixed model methods. For GWAS, we included only isolates with no mutations in known resistance genes as we were most interested in identifying novel resistance genes, and because the entire dataset contained a number of susceptible isolates with known resistance variants that would have confounded the analysis. We used pyseer v1.3.6 (52) to calculate the number of unique patterns and determine a p-value cutoff (p < 9.47e-5). Putative resistance genes were considered ones that were identified as significant by treeWAS, had more than one convergence event on the phylogeny, and >60% of all the variants in the gene were found in resistant isolates. Variants within these genes were considered putative resistance variants using the same definition as for putative resistance variants in known resistance genes. Using these requirements, we included four putative resistance variants from two putative resistance genes based on the GWAS results.

We define resistance genes as the set of all known and GWAS-identified resistance genes, and resistance variants as the set of all known and putative resistance variants in those genes.

### Identification of putative suppressor variants

We define putative suppressor variants as those in resistance genes where >60% of isolates with the variant are susceptible and contain a resistance variant.

### Comparison of isolate MICs

The significance of MIC differences between different groups of isolates was determined using two-sided Wilcox tests.

### Determination of isolate resistance group

We assigned each isolate to one of four categories (**Fig S3**):

1. <u>Resistant singletons:</u> resistant isolates that do not cluster on the phylogeny, or that cluster but contain distinct resistance variants.
2. <u>Resistant clusters:</u> resistant isolates that cluster on the phylogeny and contain the same resistance variant. Additionally, if a cluster on the phylogeny has unknown resistance variants, we also defined it as a resistant cluster.
3. <u>Susceptible revertants:</u> susceptible isolates that contain a resistance variant.
4. <u>Susceptible non-revertants:</u> susceptible isolates that do not contain a resistance variant.

For the purposes of identifying resistant clusters that include susceptible revertants, we classified an isolate as “quasi-resistant” if it contained a known resistance variant and/or was resistant. Clusters of “quasi-resistant” isolates were identified using the get_clusters function in regentrans v0.1 (https://github.com/Snitkin-Lab-Umich/regentrans) (53) with a pureness of 1. Each isolate in these clusters was then denoted as a resistant cluster or susceptible revertant isolate depending on their corresponding phenotype.

### Calculation of transmission fitness using time-scaled haplotypic density

Even though there is a clear transmission fitness cost associated with colistin resistance in the healthcare setting as evidenced by our finding that the vast majority of resistant strains do not spread, previous experimental approaches to studying the fitness of colistin-resistant CRKP have found no fitness cost to resistance (17, 54). Therefore, we define transmission fitness in the healthcare setting as the epidemic success of the strain based on time-scaled haplotypic density (THD) (24). Given a genetic distance matrix, a mutation rate, and a timescale, THD uses kernel density estimation to estimate the frequency of transmission events during that time period. We calculated the THD for each isolate with the R package thd v1.0.1 (24) using the KPNIH1 reference genome length (5,394,056 base pairs), a mutation rate of 1.03e-6 (55), the time scale parameter, and a timescale of one year. Pairwise single nucleotide variant distances were calculated with the dist.dna function in ape v5.4.1(56) (model = ‘N’, pairwise.deletion = TRUE). Prior to performing comparisons, we used the clusters identified with regentrans to remove isolates that were the only representative of their resistance group in a given cluster (i.e. all resistant singletons, single susceptible revertant isolates that were embedded in a resistant cluster, and single resistant cluster isolates that were embedded in a susceptible revertant cluster. We did this because comparisons of THD only make sense in the context of other isolates. In other words, the use of genetic context for inferring transmission fitness prevents capturing the fitness cost or benefit in these singleton isolates. Significant differences between THD of different isolates were determined using two-sided Wilcox tests.

### Calculation of previous colistin use and colistin treatment significance

We used two-sided Fisher’s exact tests to calculate the significance of the association between various groups of isolates and previous colistin use, as well as between various groups of isolates and colistin treatment.

### Data visualization

We performed all data visualization in R v4.0.2 (57) using the following packages: tidyverse v1.3.0 (58), ggtree v2.2.4 (59, 60), pheatmap v1.0.12 (61), ggplotify v0.0.5 (62), and cowplot v1.1.0 (63). Code and data corresponding to the manuscript figures can be found on GitHub (https://github.com/Snitkin-Lab-Umich/ltach-crkp-colistin-ms).

## Data Availability

Whole-genome sequences from BioProject accession no. PRJNA415194 were used.
Code and non-PHI data corresponding to the manuscript figures can be found on GitHub (https://github.com/Snitkin-Lab-Umich/ltach-crkp-colistin-ms).

https://www.ncbi.nlm.nih.gov/bioproject?term=PRJNA415194

https://github.com/Snitkin-Lab-Umich/ltach-crkp-colistin-ms

## Acknowledgements

We gratefully acknowledge Kindred Healthcare for their assistance in collecting data and isolates used in this study. We also thank Sean Muldoon for his support and guidance throughout the study. Finally, we thank the patients and staff of the long-term acute-care hospitals (LTACHs) for their gracious participation in this study, and Mary Hayden and Robert Weinstein for critical review of the manuscript.

## Funding

This research was supported by a CDC Cooperative Agreement FOA no. CK16-004-Epicenters for the Prevention of Healthcare Associated Infections and the National Institutes of Health R01 AI139240-01 and 1R01 AI148259-01. Z.L. received support from the National Science Foundation Graduate Research Fellowship Program under grant no. DGE 1256260. Any opinions, findings, and conclusions or recommendations expressed in this material are those of the authors and do not necessarily reflect the views of the National Science Foundation. The funding bodies had no role in the design of the study or collection, analysis, and interpretation of data, or in writing the manuscript.

## Supplementary Figures and Table

**Figure S1:**
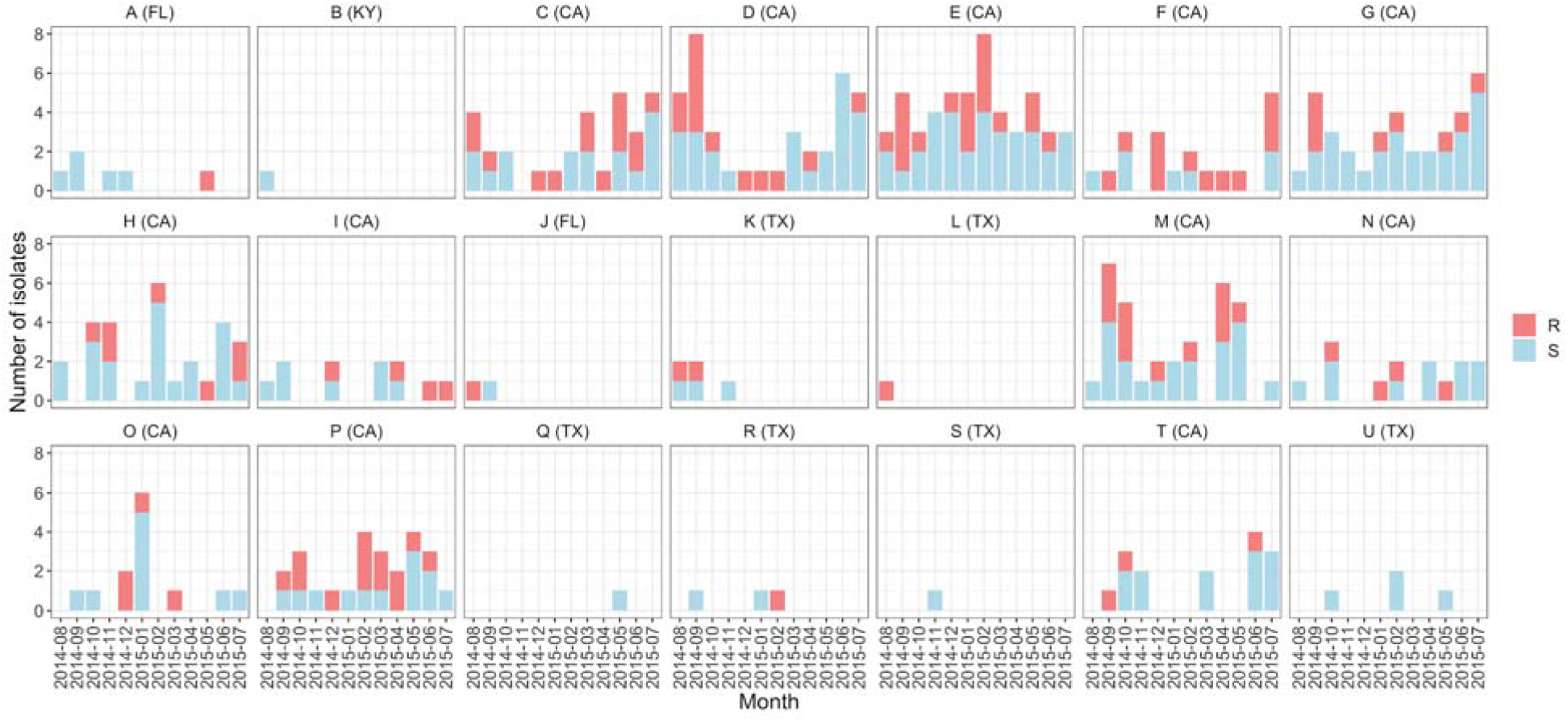
Colistin resistance occurs across time and geography. R=resistant, S=susceptible.

**Figure S2:**
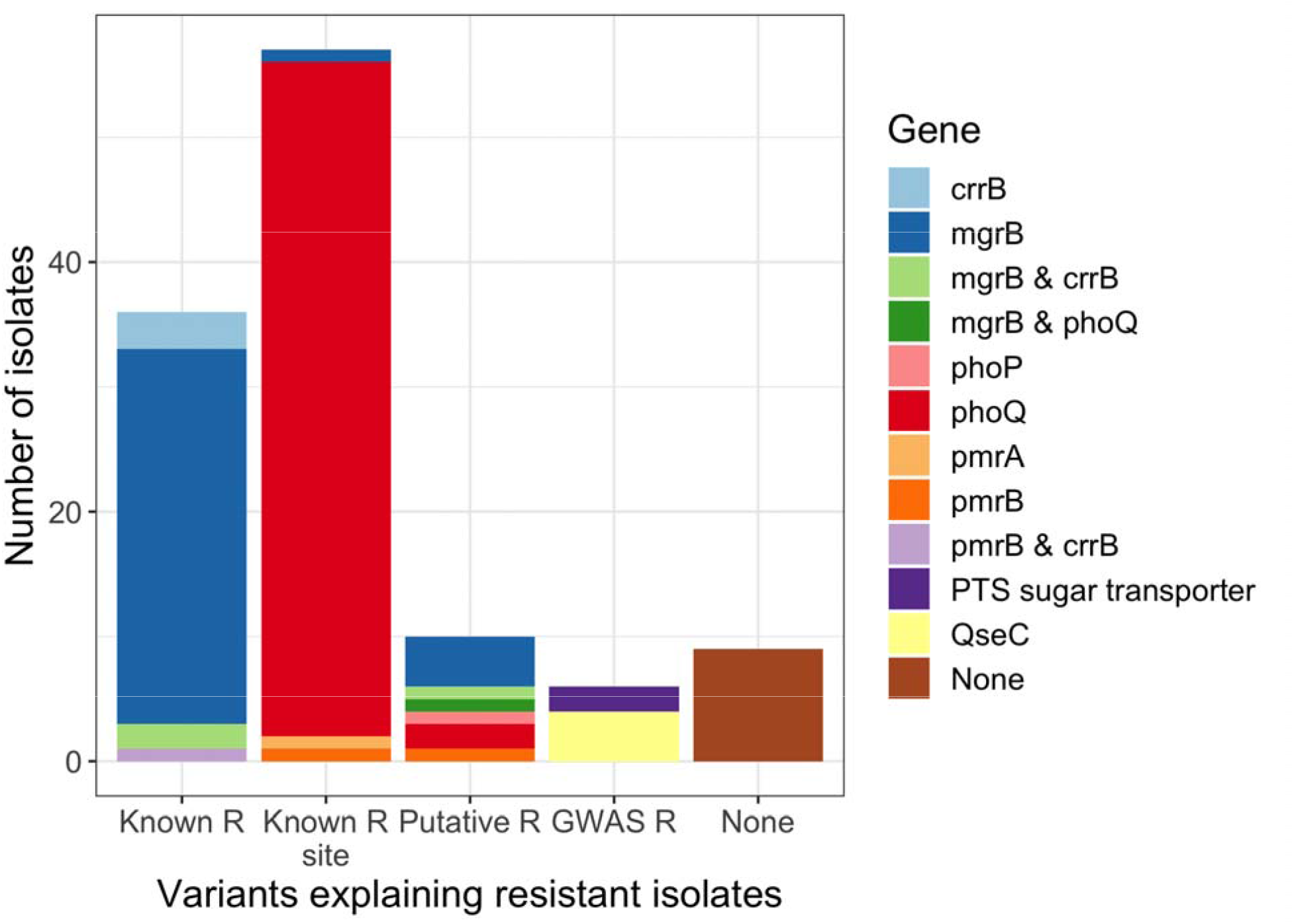
The majority of colistin resistance can be explained by variants in known resistance genes. R = resistant; GWAS = genome-wide association study. Known means the mutation or variant was previously shown to confer resistance, known site means the site was shown to confer resistance under a different amino acid change, putative and GWAS means that >60% of the variant was present in resistant isolates in canonical and GWAS genes, respectively.

**Figure S3:**
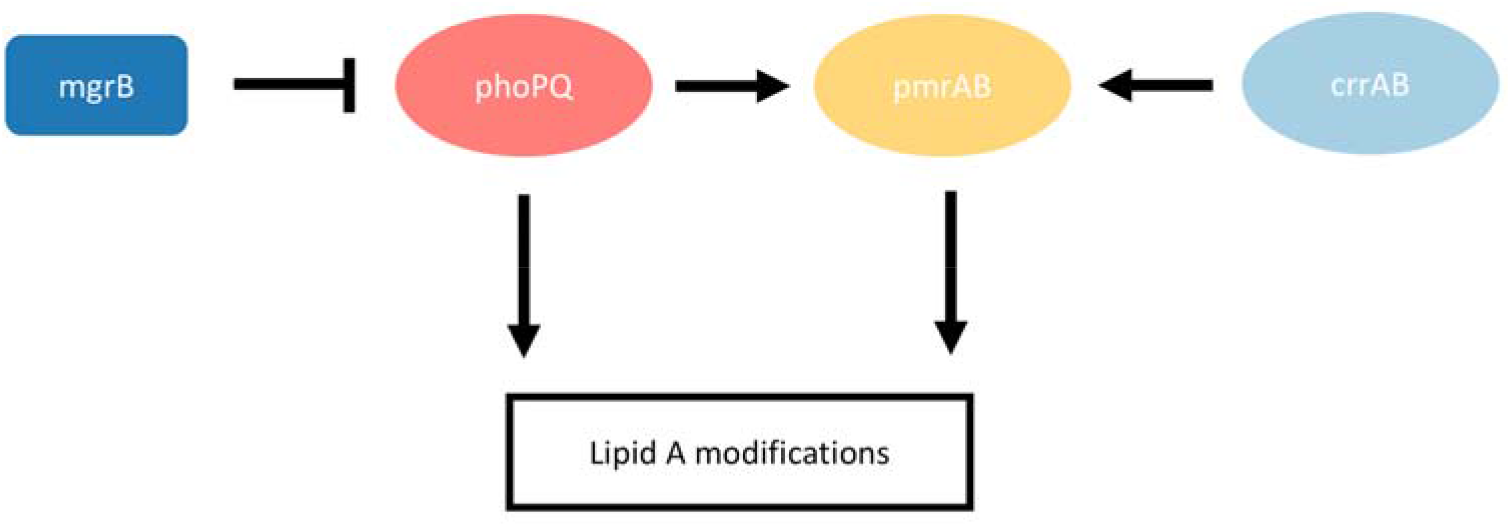
Schematic of the molecular pathway of canonical colistin resistance genes.

**Figure S4:**
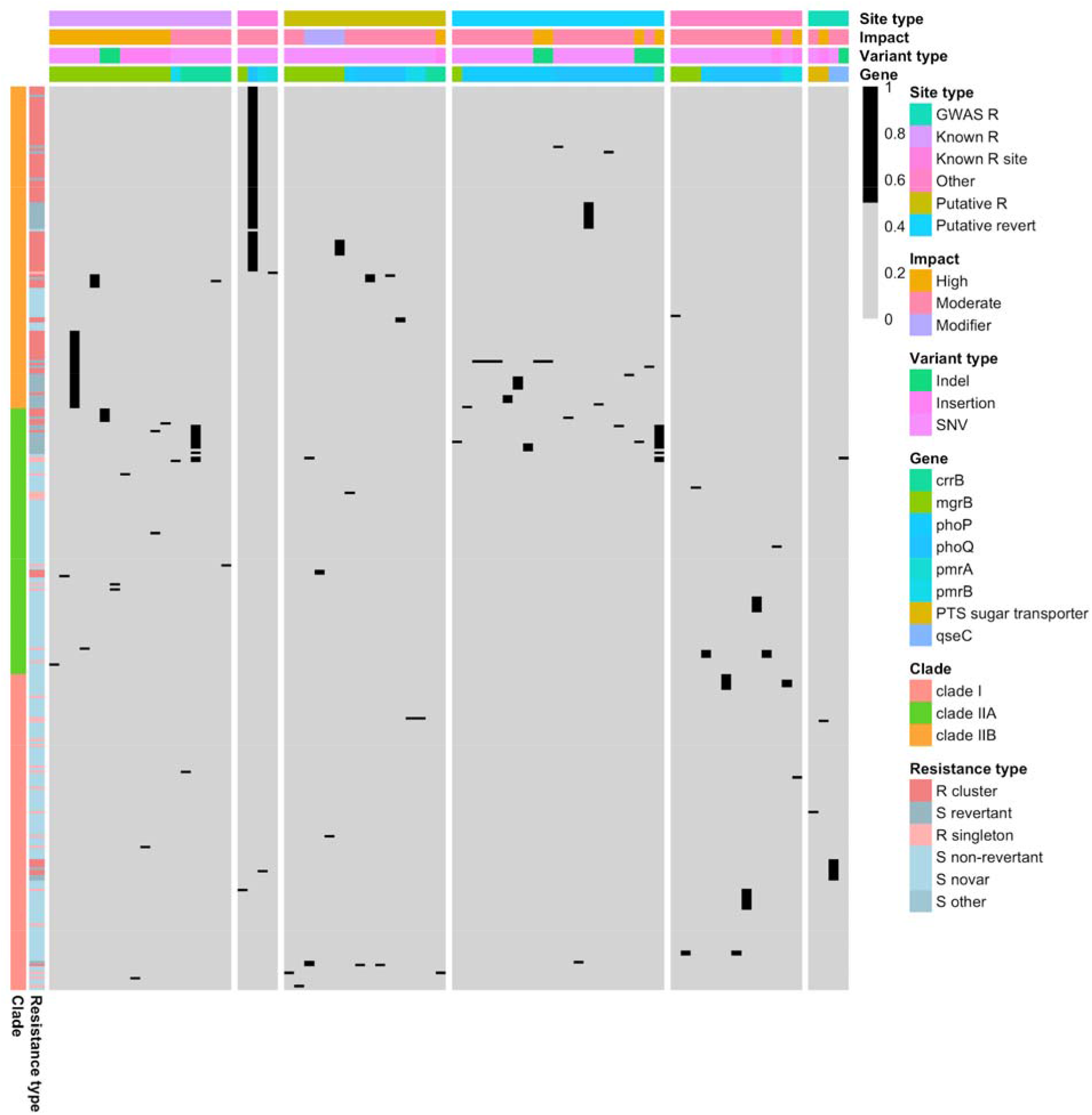
Putative resistance and suppressor variants in canonical and non-canonical resistance genes. Columns are variants and rows are isolates. All variants except the last panel are from canonical resistance genes. R=resistant, S=susceptible.

**Figure S5:**
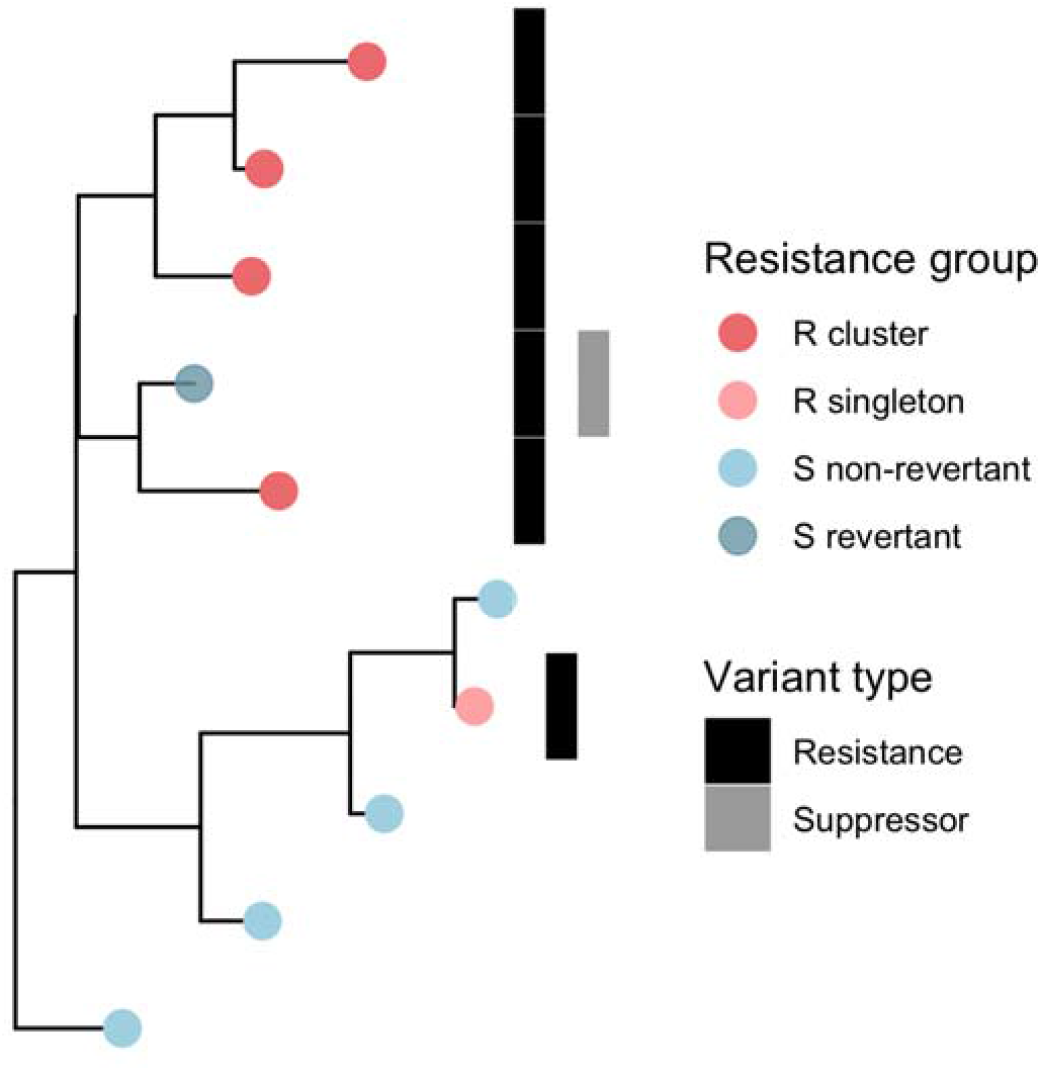
Toy example of how resistance groups are defined. Note that susceptible revertants are defined by the presence of a resistance variant, rather than a putative suppressor variant. See methods for more details. Heatmap columns are each variants found in a resistance gene. R=resistant, S=susceptible.

**Figure S6:**
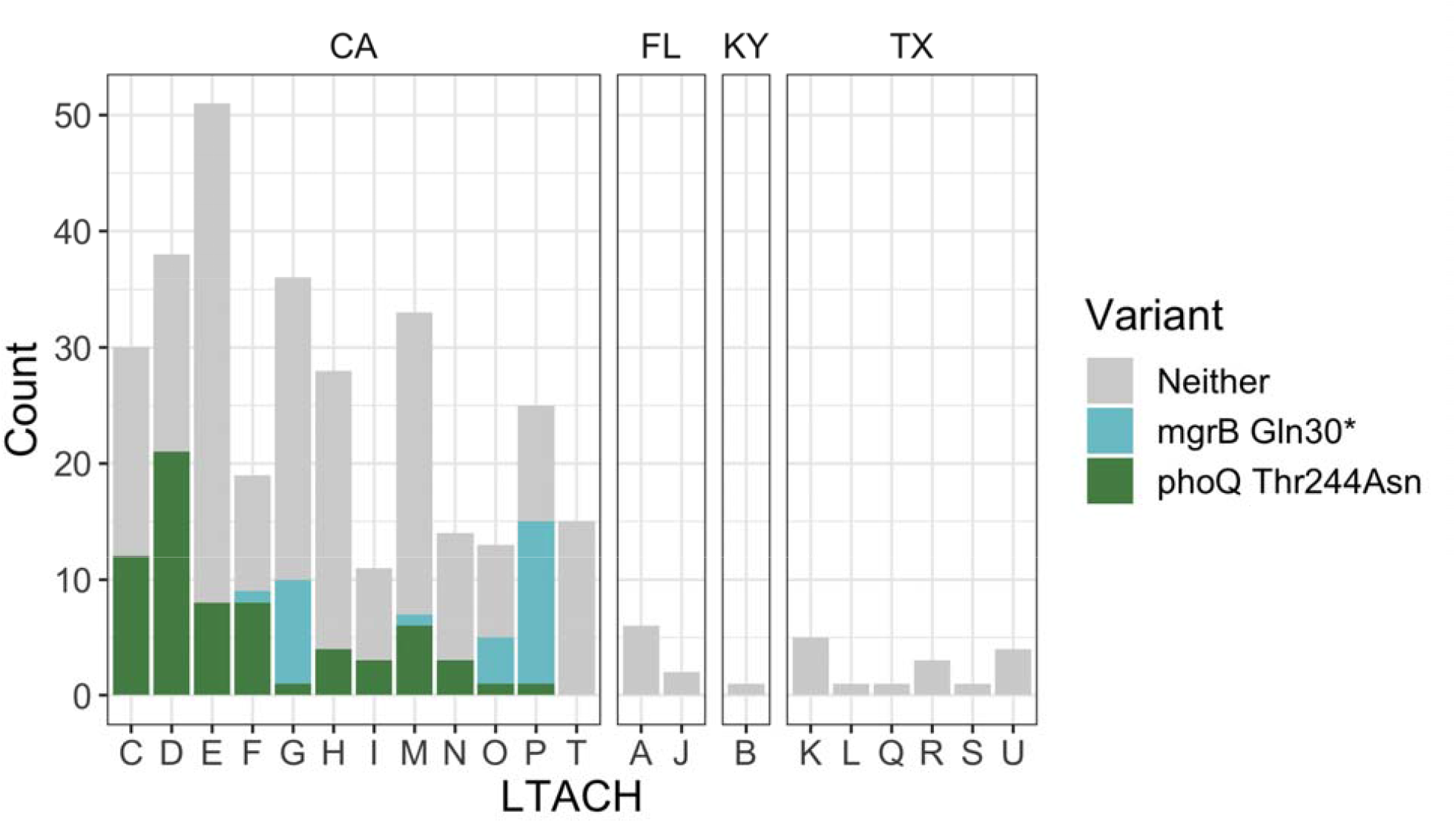
The two clonally expanded resistance variants occur across multiple LTACHs in California. 10/11 LTACHs in the Los Angeles area have the variant, as well as one LTACH in the San Diego area.

**Table S1:**
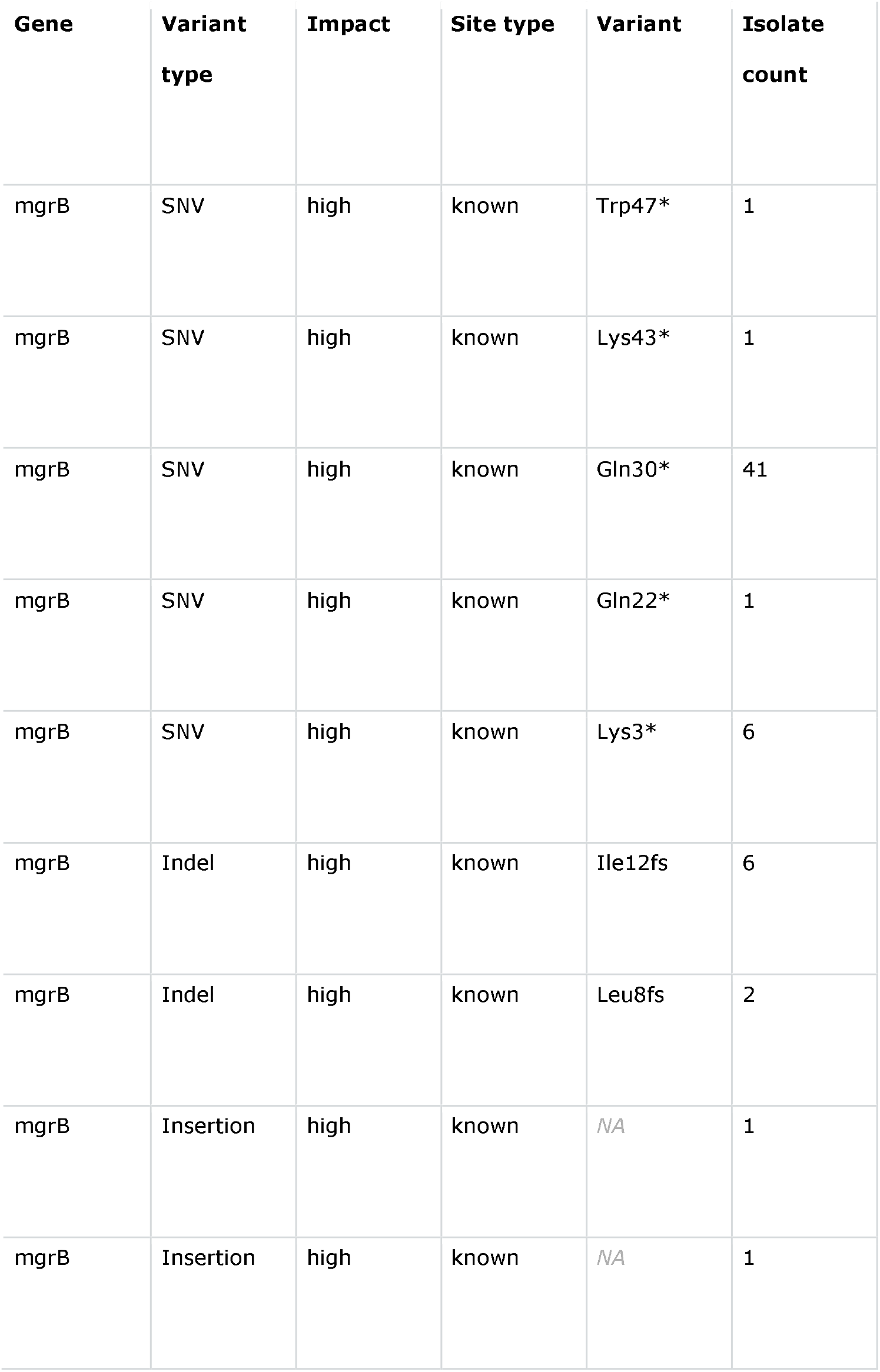
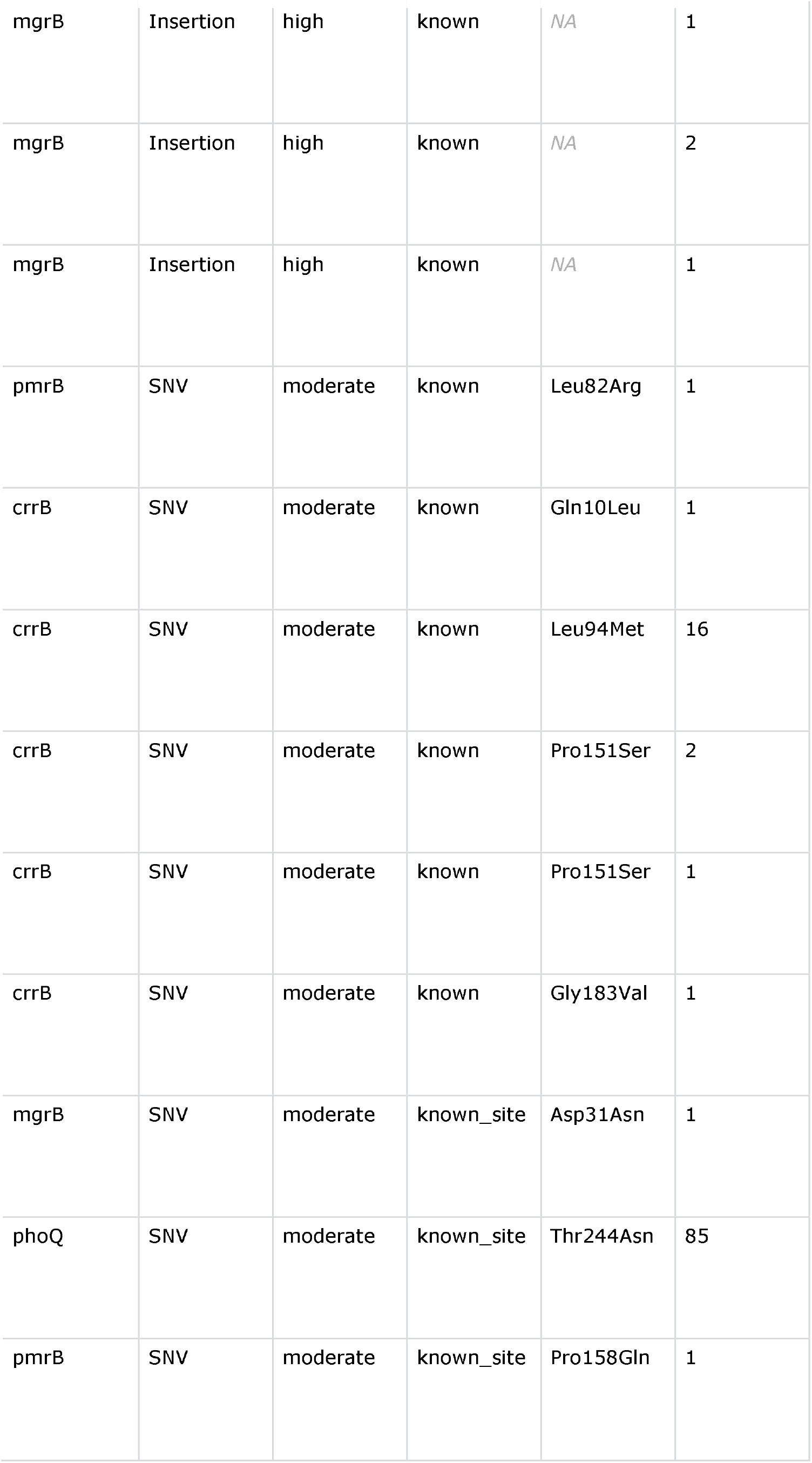
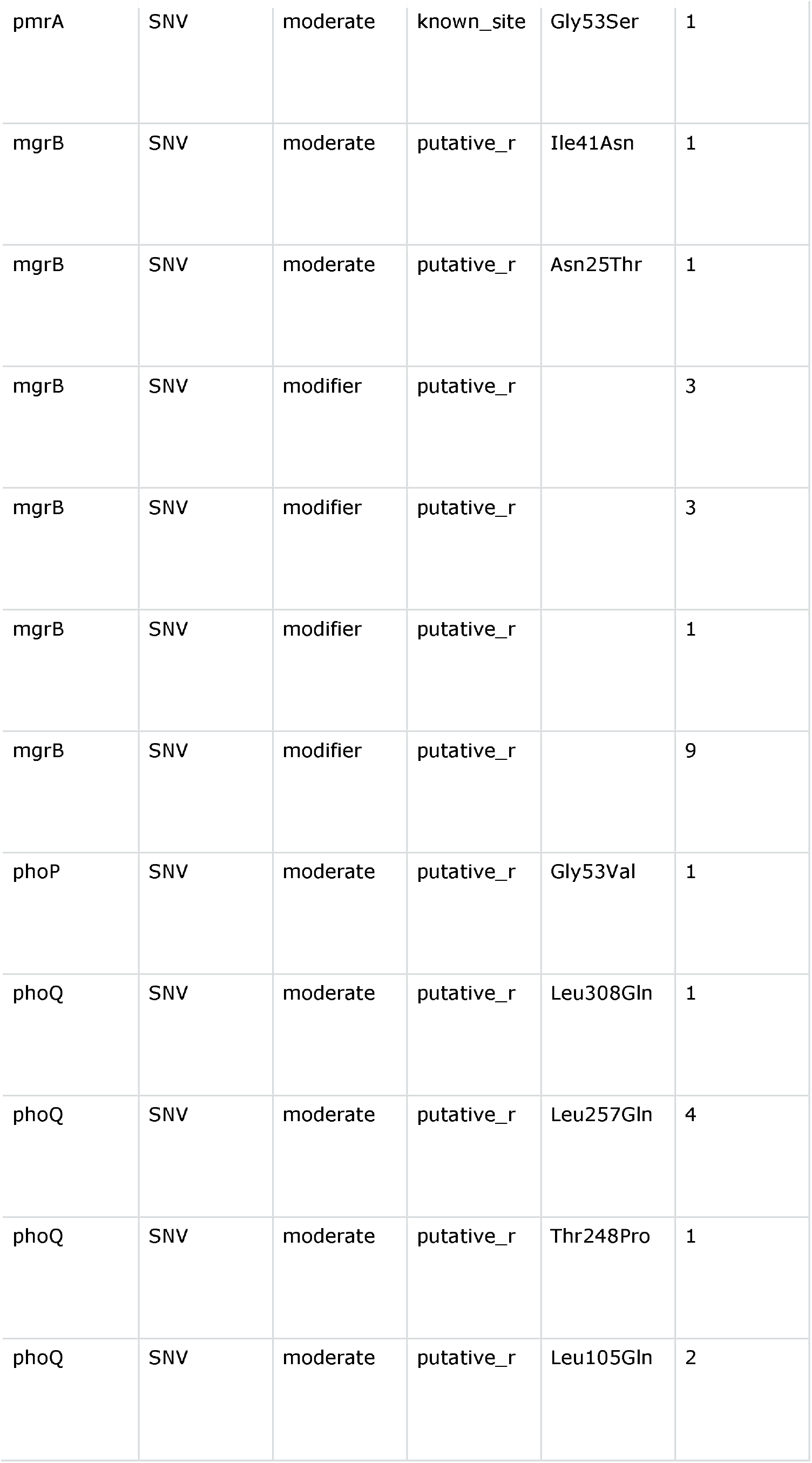
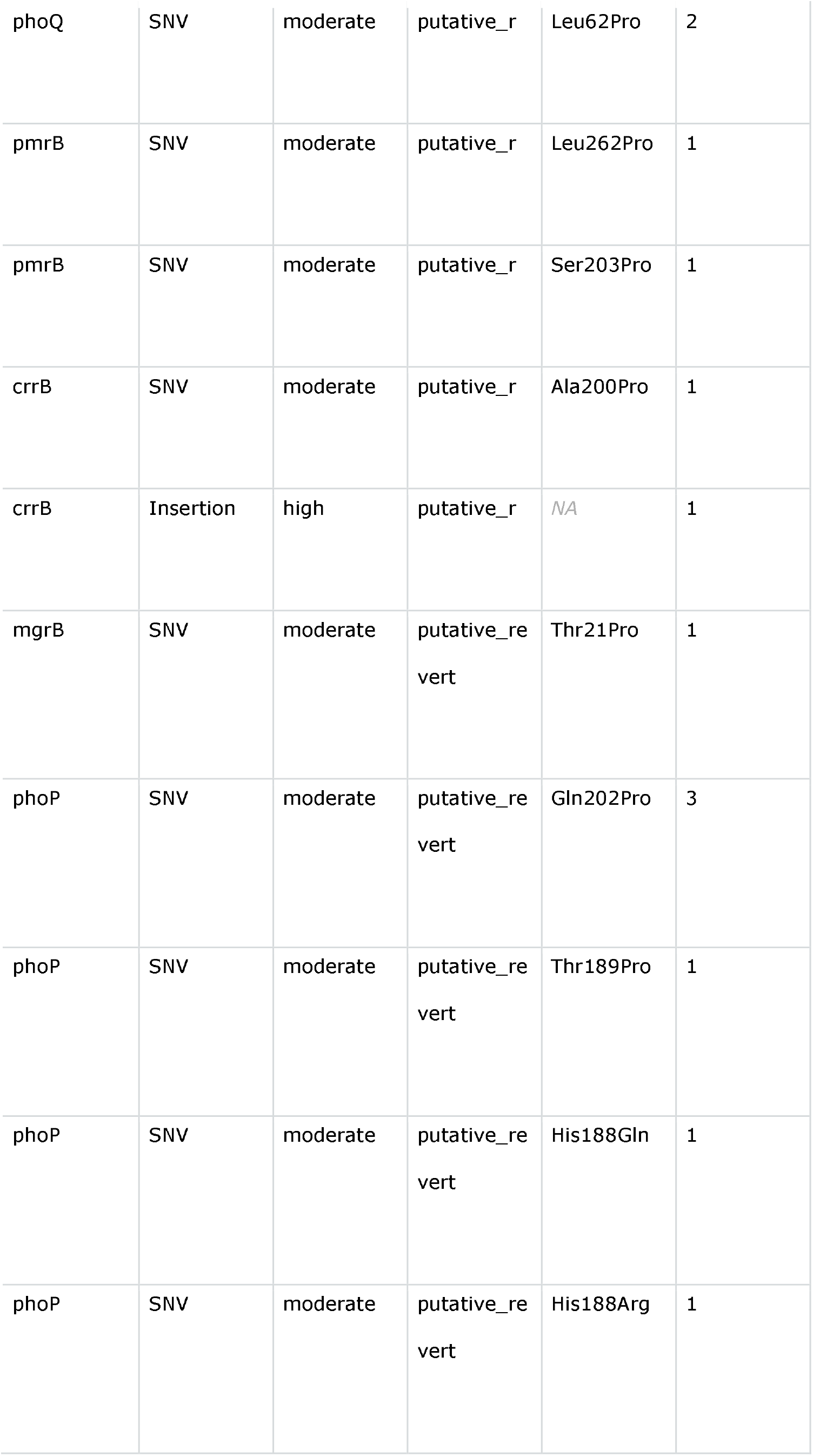
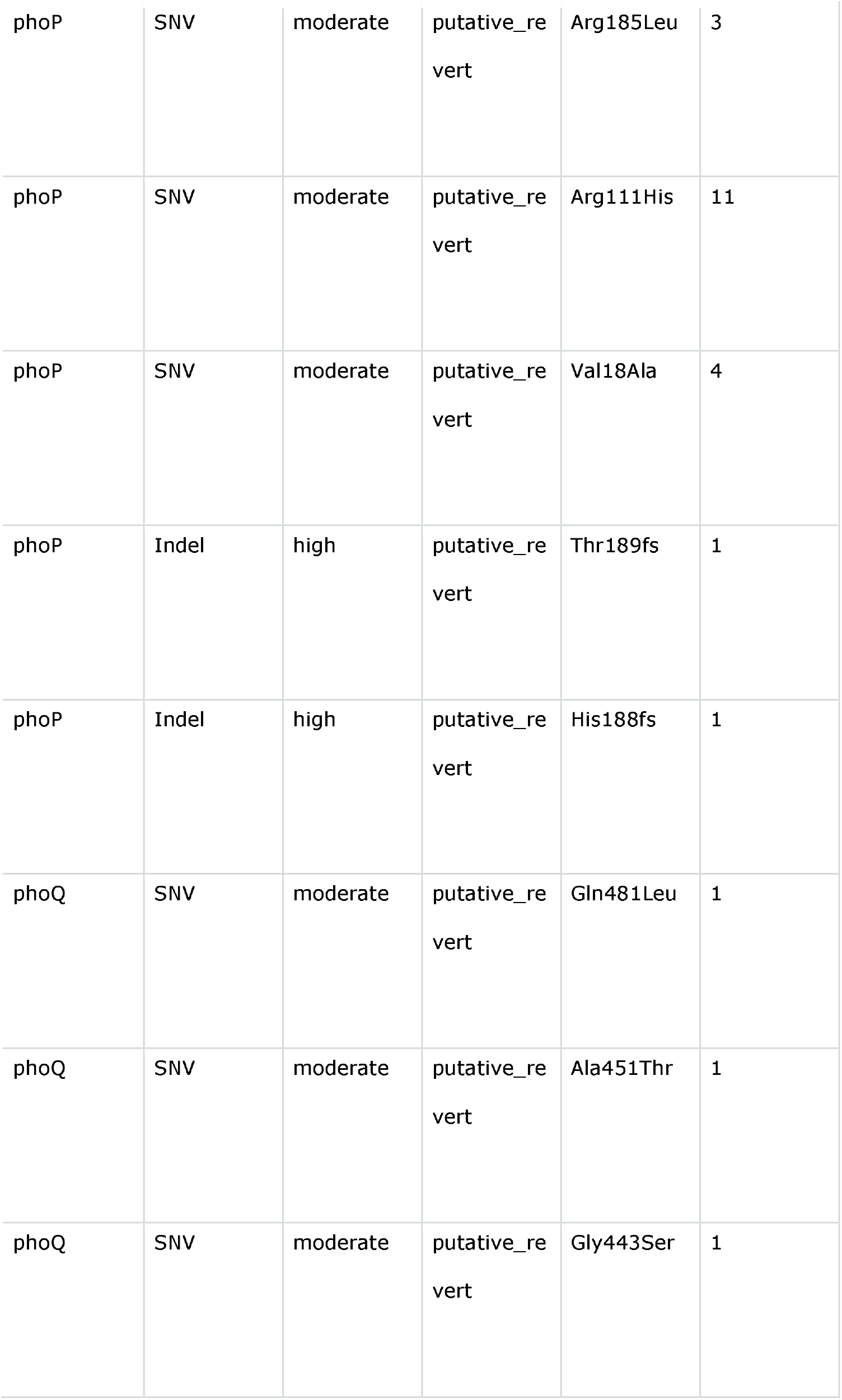
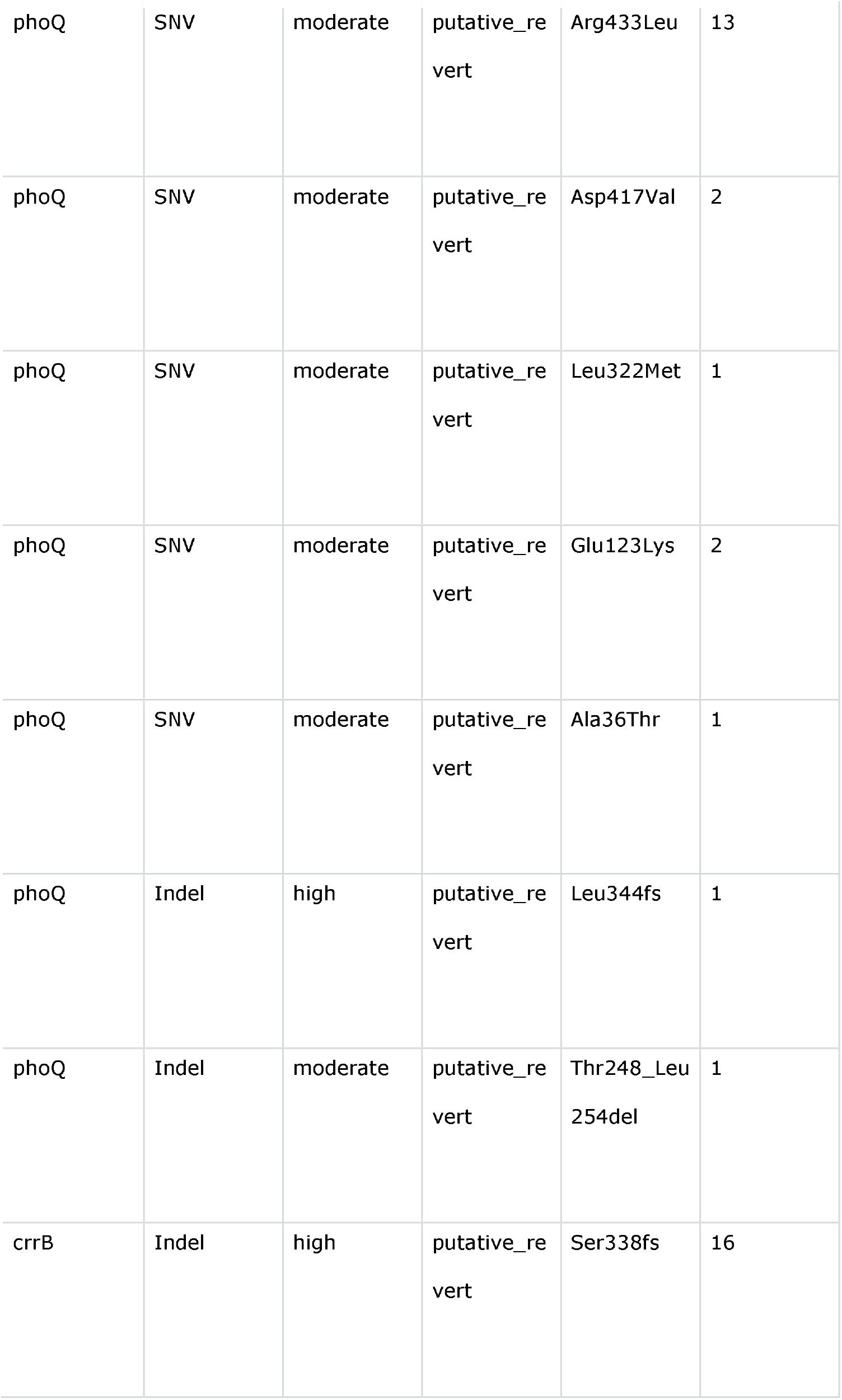
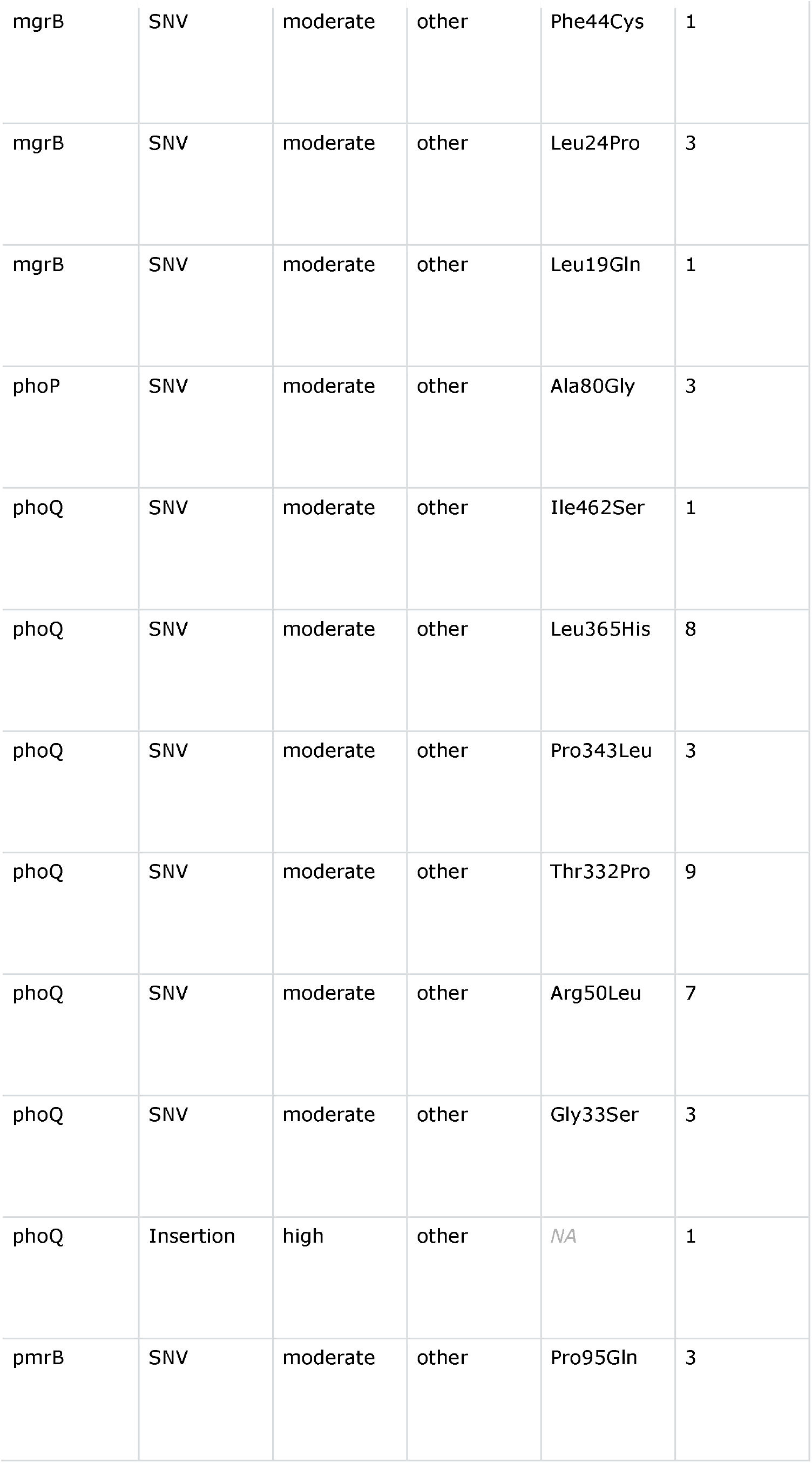
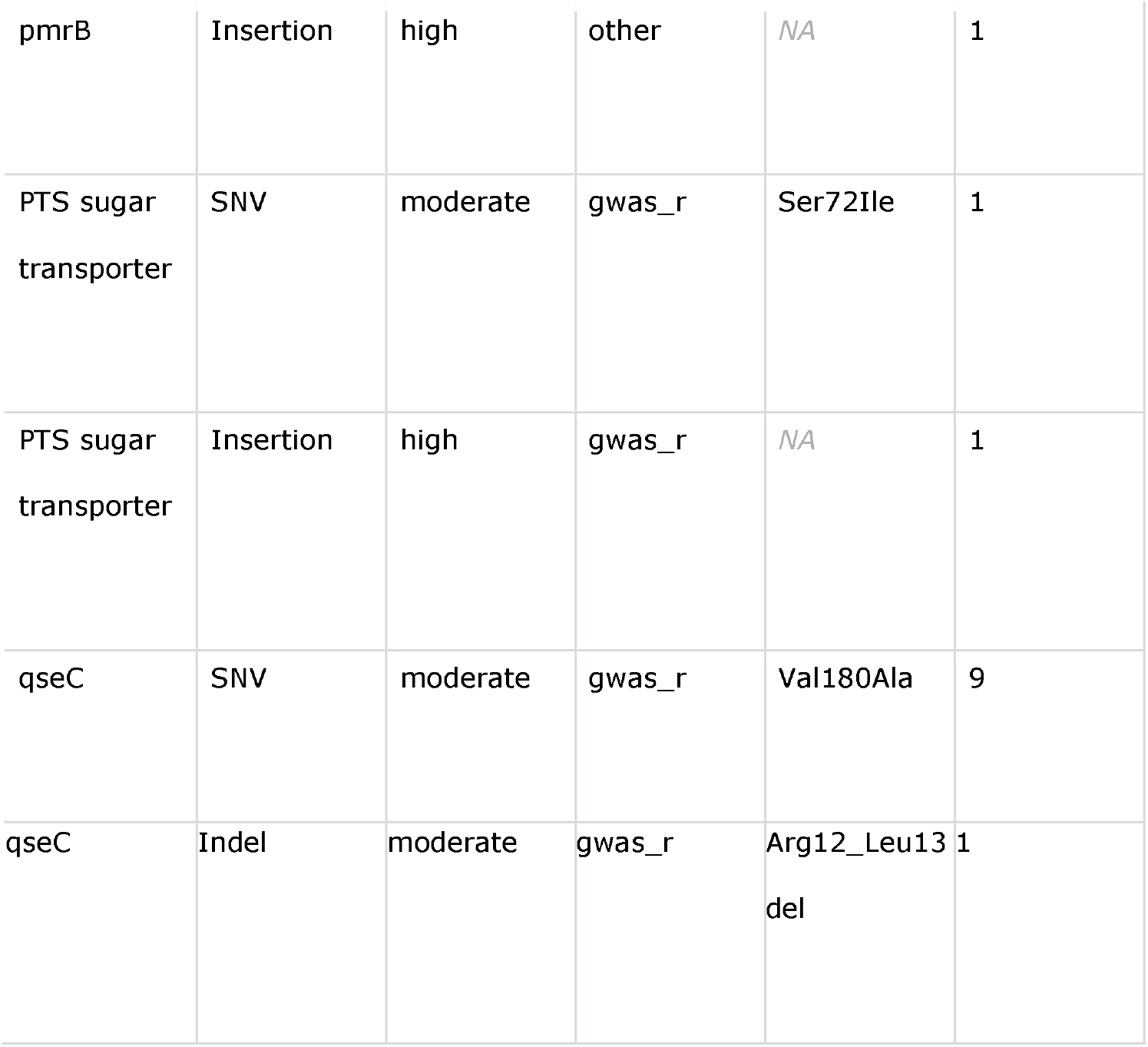
Resistance variants.

| Gene | Variant type | Impact | Site type | Variant | Isolate count |
| --- | --- | --- | --- | --- | --- |
| mgrB | SNV | high | known | Trp47* | 1 |
| mgrB | SNV | high | known | Lys43* | 1 |
| mgrB | SNV | high | known | Gln30* | 41 |
| mgrB | SNV | high | known | Gln22* | 1 |
| mgrB | SNV | high | known | Lys3* | 6 |
| mgrB | Indel | high | known | Ile12fs | 6 |
| mgrB | Indel | high | known | Leu8fs | 2 |
| mgrB | Insertion | high | known | NA | 1 |
| mgrB | Insertion | high | known | NA | 1 |
| mgrB | Insertion | high | known | NA | 1 |
| mgrB | Insertion | high | known | NA | 2 |
| mgrB | Insertion | high | known | NA | 1 |
| pmrB | SNV | moderate | known | Leu82Arg | 1 |
| crrB | SNV | moderate | known | Gln10Leu | 1 |
| crrB | SNV | moderate | known | Leu94Met | 16 |
| crrB | SNV | moderate | known | Pro151Ser | 2 |
| crrB | SNV | moderate | known | Pro151Ser | 1 |
| crrB | SNV | moderate | known | Gly183Val | 1 |
| mgrB | SNV | moderate | known_site | Asp31Asn | 1 |
| phoQ | SNV | moderate | known_site | Thr244Asn | 85 |
| pmrB | SNV | moderate | known_site | Pro158Gln | 1 |
| pmrA | SNV | moderate | known_site | Gly53Ser | 1 |
| mgrB | SNV | moderate | putative_r | Ile41Asn | 1 |
| mgrB | SNV | moderate | putative_r | Asn25Thr | 1 |
| mgrB | SNV | modifier | putative_r |  | 3 |
| mgrB | SNV | modifier | putative_r |  | 3 |
| mgrB | SNV | modifier | putative_r |  | 1 |
| mgrB | SNV | modifier | putative_r |  | 9 |
| phoP | SNV | moderate | putative_r | Gly53Val | 1 |
| phoQ | SNV | moderate | putative_r | Leu308Gln | 1 |
| phoQ | SNV | moderate | putative_r | Leu257Gln | 4 |
| phoQ | SNV | moderate | putative_r | Thr248Pro | 1 |
| phoQ | SNV | moderate | putative_r | Leu105Gln | 2 |
| phoQ | SNV | moderate | putative_r | Leu62Pro | 2 |
| pmrB | SNV | moderate | putative_r | Leu262Pro | 1 |
| pmrB | SNV | moderate | putative_r | Ser203Pro | 1 |
| crrB | SNV | moderate | putative_r | Ala200Pro | 1 |
| crrB | Insertion | high | putative_r | NA | 1 |
| mgrB | SNV | moderate | putative_re<br>vert | Thr21Pro | 1 |
| phoP | SNV | moderate | putative_re<br>vert | Gln202Pro | 3 |
| phoP | SNV | moderate | putative_re<br>vert | Thr189Pro | 1 |
| phoP | SNV | moderate | putative_re<br>vert | His188Gln | 1 |
| phoP | SNV | moderate | putative_re<br>vert | His188Arg | 1 |
| phoP | SNV | moderate | putative_re<br>vert | Arg185Leu | 3 |
| phoP | SNV | moderate | putative_re<br>vert | Arg111His | 11 |
| phoP | SNV | moderate | putative_re<br>vert | Val18Ala | 4 |
| phoP | Indel | high | putative_re<br>vert | Thr189fs | 1 |
| phoP | Indel | high | putative_re<br>vert | His188fs | 1 |
| phoQ | SNV | moderate | putative_re<br>vert | Gln481Leu | 1 |
| phoQ | SNV | moderate | putative_re<br>vert | Ala451Thr | 1 |
| phoQ | SNV | moderate | putative_re<br>vert | Gly443Ser | 1 |
| phoQ | SNV | moderate | putative_re<br>vert | Arg433Leu | 13 |
| phoQ | SNV | moderate | putative_re<br>vert | Asp417Val | 2 |
| phoQ | SNV | moderate | putative_re<br>vert | Leu322Met | 1 |
| phoQ | SNV | moderate | putative_re<br>vert | Glu123Lys | 2 |
| phoQ | SNV | moderate | putative_re<br>vert | Ala36Thr | 1 |
| phoQ | Indel | high | putative_re<br>vert | Leu344fs | 1 |
| phoQ | Indel | moderate | putative_re<br>vert | Thr248_Leu<br>254del | 1 |
| crrB | Indel | high | putative_re<br>vert | Ser338fs | 16 |
| mgrB | SNV | moderate | other | Phe44Cys | 1 |
| mgrB | SNV | moderate | other | Leu24Pro | 3 |
| mgrB | SNV | moderate | other | Leu19Gln | 1 |
| phoP | SNV | moderate | other | Ala80Gly | 3 |
| phoQ | SNV | moderate | other | Ile462Ser | 1 |
| phoQ | SNV | moderate | other | Leu365His | 8 |
| phoQ | SNV | moderate | other | Pro343Leu | 3 |
| phoQ | SNV | moderate | other | Thr332Pro | 9 |
| phoQ | SNV | moderate | other | Arg50Leu | 7 |
| phoQ | SNV | moderate | other | Gly33Ser | 3 |
| phoQ | Insertion | high | other | *NA* | 1 |
| pmrB | SNV | moderate | other | Pro95Gln | 3 |
| pmrB | Insertion | high | other | NA | 1 |
| PTS sugar transporter | SNV | moderate | gwas_r | Ser72Ile | 1 |
| PTS sugar transporter | Insertion | high | gwas_r | NA | 1 |
| qseC | SNV | moderate | gwas_r | Val180Ala | 9 |
| qseC | Indel | moderate | gwas_r | Arg12_Leu13 del | 1 |

## Notes

### Author Declarations

No new data used. The original study was reviewed and approved by the Institutional Review Board of the University of Pennsylvania with a waiver of informed consent.

### Summary of Updates

Main text, particularly the results, reworded for clarity. Minor changes to figures.

